# A methylome-wide association study of major depression with out-of-sample case-control classification and trans-ancestry comparison

**DOI:** 10.1101/2023.10.27.23297630

**Authors:** Xueyi Shen, Miruna Barbu, Doretta Caramaschi, Ryan Arathimos, Darina Czamara, Friederike S. David, Anna Dearman, Evelyn Dilkes, Marisol Herrera-Rivero, Floris Huider, Luise Kühn, Kuan-Chen Lu, Teemu Palviainen, Alicia Marie Schowe, Gemma Shireby, Antoine Weihs, Chloe C. Y. Wong, Eleanor Davyson, Hannah Casey, Mark J Adams, Antje-Kathrin Allgaier, Michael Barber, Joe Burrage, Avshalom Caspi, Ricardo Costeira, Erin C. Dunn, Lisa Feldmann, Josef Frank, Franz Joseph Freisleder, Danni A. Gadd, Ellen Greimel, Eilis Hannon, Sarah E Harris, Georg Homuth, David M. Howard, Stella Iurato, Tellervo Korhonen, Tzu-Pin Lu, Nicholas G Martin, Jade Martins, Edel McDermott, Susanne Meinert, Pau Navarro, Miina Ollikainen, Verena Pehl, Charlotte Piechaczek, Aline D. Scherff, Frederike Stein, Fabian Streit, Alexander Teumer, Henry Völzke, Jenny van Dongen, Rosie M. Walker, Natan Yusupov, Louise Arseneault, Jordana T. Bell, Klaus Berger, Elisabeth Binder, Dorret I. Boomsma, Simon R Cox, Udo Dannlowski, Kathryn L. Evans, Helen L. Fisher, Andreas J. Forstner, Hans J. Grabe, Jaakko Kaprio, Tilo Kircher, Johannes Kopf-Beck, Meena Kumari, Po-Hsiu Kuo, Qingqin S Li, Terrie E. Moffitt, Hugh Mulcahy, Therese M. Murphy, Gerd Schulte-Körne, Jonathan Mill, Cathryn M. Lewis, BeCOME Working Group, OPTIMA Working Group, PGC MDD Working Group, Naomi R Wray, Andrew M McIntosh

**Author notes:** Joint contribution (Xueyi Shen, Miruna Barbu). Corresponding authors, Dr Xueyi Shen Address: University of Edinburgh, Royal Edinburgh Hospital, Morningside Park Edinburgh EH10 5HF, United Kingdom, Prof Andrew M McIntosh, Address: University of Edinburgh, Royal Edinburgh Hospital, Morningside Park Edinburgh EH10 5HF United Kingdom.

## Abstract

Major Depression (MD) is a leading cause of global disease burden, and both experimental and population-based studies suggest that differences in DNA methylation (DNAm) may be associated with the condition. However, previous DNAm studies have not so far been widely replicated, suggesting a need for larger meta-analysis studies. In the present study, the Psychiatric Genomics Consortium Major Depressive Disorder working group conducted a meta-analysis of methylome-wide association analysis (MWAS) for life-time MD across 18 studies of 24,754 European-ancestry participants (5,443 MD cases) and an East Asian sample (243 cases, 1846 controls). We identified fifteen CpG sites associated with lifetime MD with methylome-wide significance (p < 6.42×10^-8^). Top CpG effect sizes in European ancestries were positively correlated with those from an independent East Asian MWAS (r = 0.482 and p = 0.068 for significant CpG sites, r = 0.261 and p = 0.009 for the top 100 CpG sites). Methylation score (MS) created using the MWAS summary statistics was significantly associated with MD status in an out-of-sample classification analysis (β = 0.122, p = 0.005, AUC = 0.53). MS was also associated with five inflammatory markers, with the strongest association found with Tumor Necrosis Factor Beta (β=-0.154, p=1.5×10^-5^). Mendelian randomisation (MR) analysis demonstrated that 23 CpG sites were potentially causally associated with MD and six of those were replicated in an independent mQTL dataset (Wald’s ratio test, absolute β ranged from 0.056 to 0.932, p ranged from 7×10^-3^ to 4.58×10^-6^). CpG sites located in the Major Histocompatibility complex (MHC) region showed the strongest evidence from MR analysis of being associated with MD. Our study provides evidence that variations in DNA methylation are associated with MD, and further evidence supporting involvement of the immune system. Larger sample sizes in diverse ancestries are likely to reveal replicable associations to improve mechanistic inferences with the potential to inform molecular target identification.

## Introduction

Major depression (MD) is a common psychiatric disorder arising from a complex combination of genetic and environmental factors that include lifestyle factors such as physical activity, smoking, alcohol consumption, and body mass index (BMI)^1–3^. The heritability of MD estimated from twin studies is 37%^4^, and polygenic risk scores (PRS) trained on genome-wide-association (GWAS) currently explain 1.5-3.2% of the variance in MD^3^.

DNA methylation (DNAm) is one of the most studied epigenetic processes and is influenced by both genetic and environmental factors^5^. DNAm is dynamic and is associated with changes in environmental and lifestyle factors, including smoking^6^, alcohol^7^, and BMI^8^, all factors that are implicated in MD^3^. A recent study^9^ identified associations between methylation scores (MS), calculated using methylome-wide association study (MWAS) summary statistics for several relevant lifestyle factors and MD^10^. These environmental MS measures were able to capture additional variation associated with MD when added to direct lifestyle measures, and it is thought this may be due to their ability to act as an archive of environmental exposure.

There is growing evidence from MWAS that DNAm measured in whole blood may be associated with MD. Jovanova et al. (2018) looked at 11 cohorts comprising 11,256 participants of European and African ancestry and identified 3 cytosine-phosphate-guanine (CpG) sites associated with depressive symptoms, which were annotated to genes implicated in axon guidance^11^. Starnawska et al. (2019) investigated depressive symptomatology in a sample of 724 monozygotic twins, with top findings annotated to genes previously implicated in depression^12^. Finally, Huls et al. (2020) identified DNAm associations with MD in dorsal lateral prefrontal cortex samples from N=608 participants and uncovered CpGs annotated to several genes that are relevant to MD^13^. In addition, MS calculated using penalised regression models have previously been utilised to investigate MD^14^. We recently used lasso regression to calculate an MD MS in N=1,780 participants and found that the scores explain approximately 1.75% of the variance in liability to life-time MD, acting additively with PRS^14^. Findings from these studies have been somewhat inconsistent regarding the specific associations. Larger MWAS studies may provide more reliable estimates of the differences in DNAm between MD cases and controls, and in doing so, bring insight to the molecular mechanisms associated with MD.

Given previous evidence highlighting the role of DNAm in MD, we conducted an MD MWAS meta-analysis using DNA extracted from whole blood in 18 cohorts comprising 24,754 individuals (5,443 cases) of European ancestry. We sought to identify whether differences in DNAm were a potentially cause or consequence of MD using a two-sample Mendelian Randomisation framework. Further, we trained a DNAm classifier of MD status from our summary statistics, and assessed whether it could classify MD case-control status in an independent testing sample and its association with the abundance of inflammatory protein markers. Finally, we assessed whether significant effect sizes were positively associated with those in an independent East Asian sample.

## Methods

### Cohort information

#### Participants

A total of 24,754 European-ancestry participants (5,443 MD cases) from 18 studies were included in the meta-analysis. The mean age of participants in each study ranged from 15 to 59 years. Details for each individual study can be found in Table 1 and Supplementary Methods. Written consents were obtained from all participants.

**Table 1.**
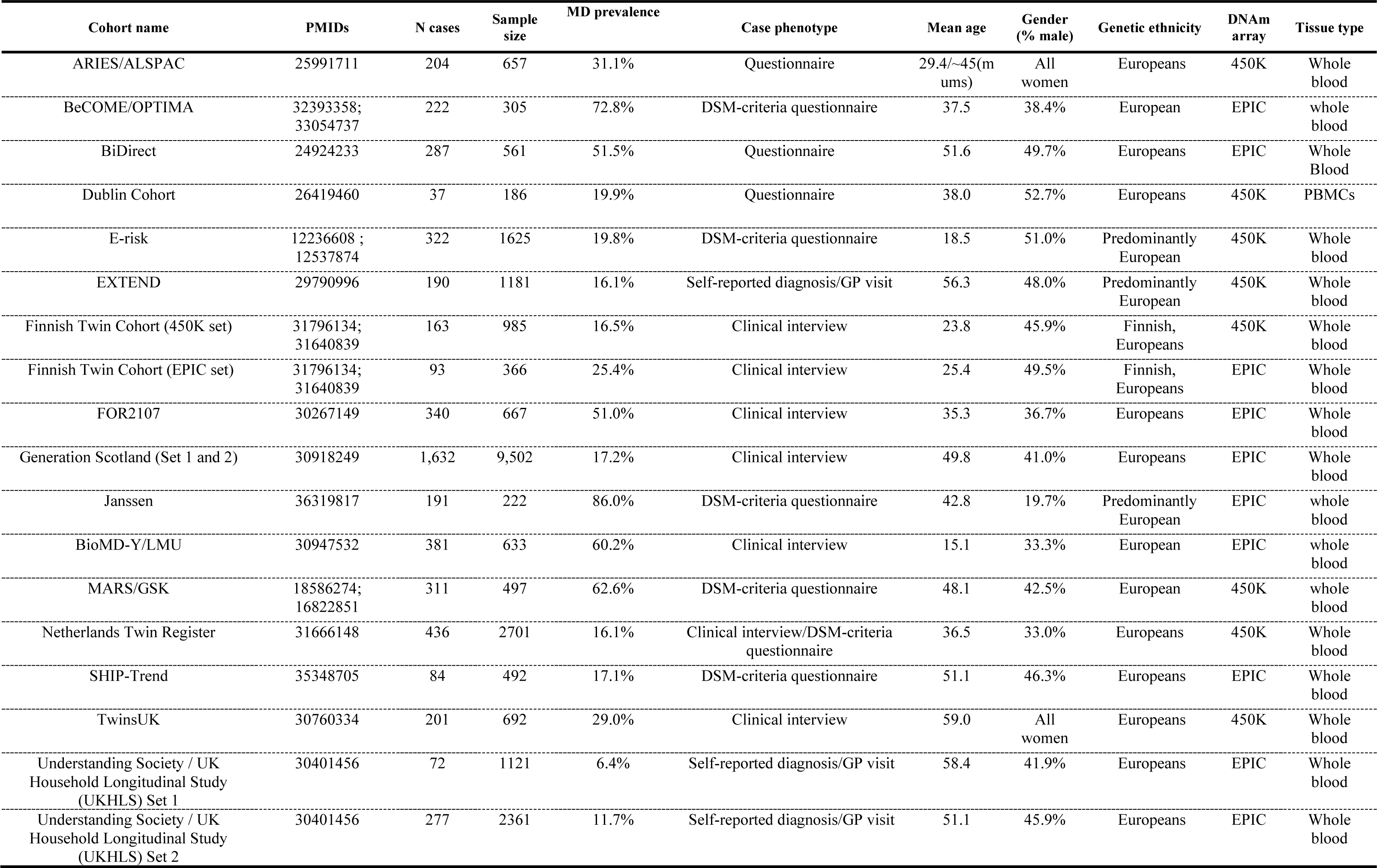
Information for cohorts that participated in the methylome-wide meta-analysis.

#### DNAm data preparation and quality check

DNAm data were obtained from DNA extracted from whole blood. Eight studies used the Infinium Human Methylation 450 (450K) BeadChip array (Illumina Inc., number of CpG sites ranged from 275,868 to 438,752 after quality check) and the other ten studies used the Illumina Infinium Methylation EPIC array (Illumina Inc., number of CpG sites ranged from 673,085 to 809,447 after quality check). Quality checks and data normalisation were conducted by each individual study team. Details are provided in the protocol papers for each individual study (Table 1 and Supplementary Methods). In brief, the majority of studies used functional normalisation for methylation data pre-processing, unless stated otherwise in the Supplementary Methods^15^. Similar quality check procedures were used, including removing probes with outlying signal intensity, bead count and detection p-values, removing participants with mismatched sex prediction from DNA methylation data and removing cross-hybridising probes that map to common genetic variants (at MAF > 0.05) and polymorphic probes^15^. M-values were used for the association analysis^16^.

#### Diagnosis of Major Depression (MD)

Life-time diagnosis for MD were derived based on structured clinical interview or self-reported symptoms. Those studies that derived diagnoses of MD based on structural clinical interviews used criteria from The Diagnostic and Statistical Manual of Mental Disorders, Fifth/Fourth Edition (DSM-5/DSM-4)^17^. Self-declared MD was based on depressive symptoms lasting for more than two weeks and help-seeking due to depression. Studies that derived MD diagnosis based on multiple time points defined cases as those who experienced any depressive episodes during their lifetime, and controls were those who did not declare MD throughout. A total of 7 studies defined MD cases using structural clinical interview (N cases = 3,246), 5 studies used DSM-criteria questionnaires (N cases = 1,130), 3 studies used self-administered questionnaires for depressive symptoms (N cases = 528), and 3 studies defined MD cases based on self-declared visits to general practitioner (N cases = 539). Details for MD diagnosis for each cohort can be found in Table 1 and the Supplementary Materials, and Methods sections.

Additional exclusion criteria per study are stated in the Supplementary Methods.

#### Association analysis

Linear regression was used to test the association between DNA methylation (M-values, dependent variable) and MD diagnosis (independent variable) using a pipeline available at the URL: https://github.com/psychiatric-genomics-consortium/mdd-mwas. Those cohorts that used their own specific pipelines were specified in the Supplementary Methods. The pipeline uses the R package ‘limma’ for linear regression on large omic data^18^. Three models representing increasingly rigorous correction for potential confounders were estimated. The simplest model contained age, sex, batch, the first 20 methylation principal components (PCs) or surrogate variables (SVs)^19^ based on the study protocol for each individual cohort, and white-blood cell proportions estimated from DNA methylation data of CD8+T, CD4+T, natural killer cells, B cells and granulocytes^19^. The AHRR (aryl hydrocarbon receptor repressor)-adjusted model had an additional covariate that adjusted for smoking status by including the M-values for the AHRR probe (cg05575921), due to its known accuracy in predicting smoking^20^ and its consistency and availability in all studies. Finally, a third model with additional covariates (referred to as the ‘complex model’) was fitted that contained body mass index (BMI) and alcohol consumption in addition to all the other covariates included in the previous models.

Results for the AHRR-adjusted model (referred to as the ‘main model’) are reported as the main findings. Standardised Cohen’s d between MD cases versus controls were reported as effect sizes.

#### Meta analysis

Meta analysis was conducted using METAL (version 2011)^21^ in a two-stage process. First, meta-analysis was performed on studies that used 450K and EPIC arrays separately, due to the difference of CpG sites available for each array (Figure 1).

**Figure 1.**
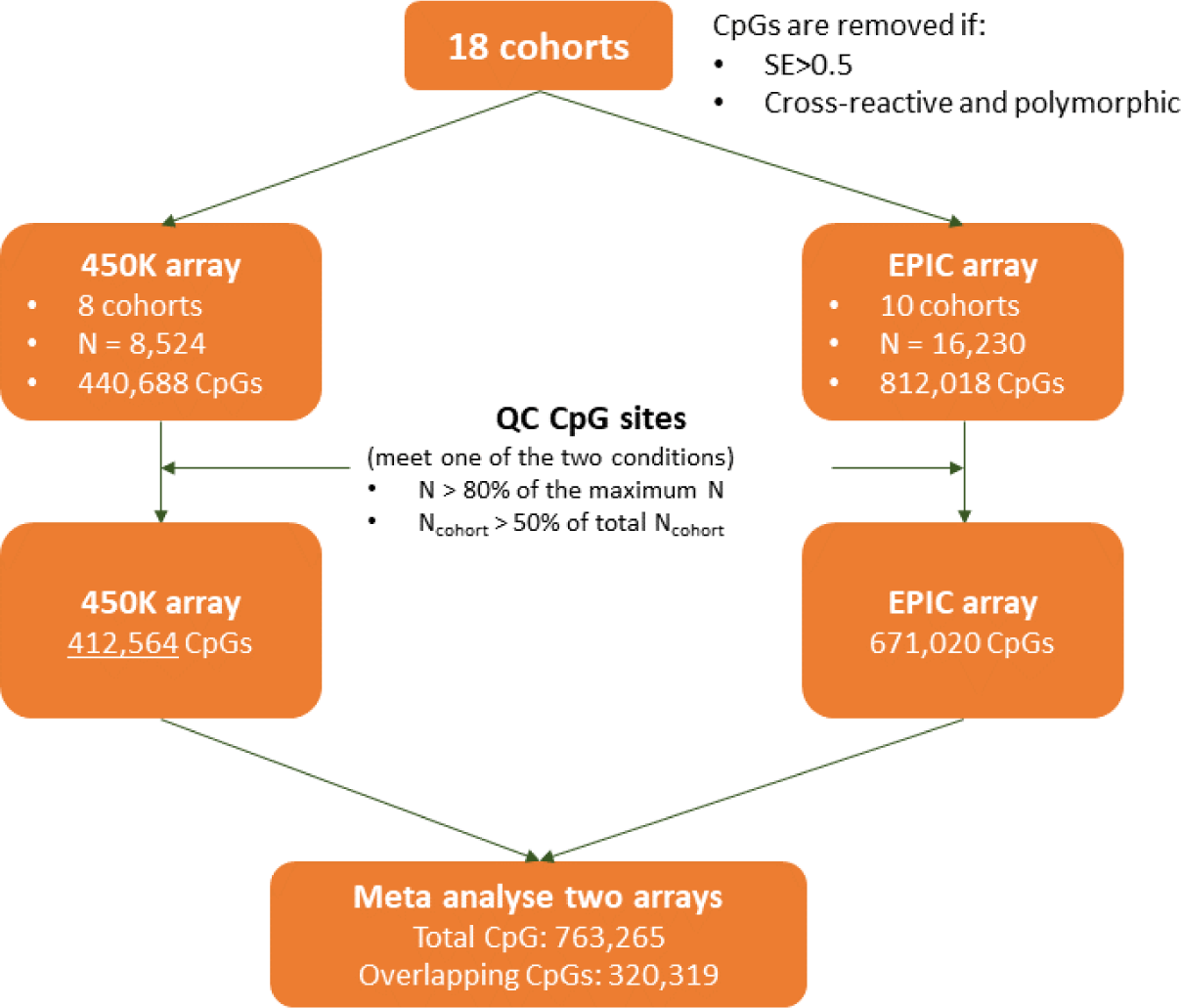
Workflow of meta-analysis.

Those CpG sites that were either available for more than half of the studies using the given array or had a total sample size over 80% of the max sample size were kept for further analysis. CpG sites with excessive standard errors (SE>0.5, see Supplementary Figure 1) were removed from analysis.

Second, the summary statistics for 450K and EPIC array were meta-analysed. A fixed-effect inverse-variance model was used without genomic control correction. P-values were Bonferroni corrected (p-value threshold = 6.55×10^-8^ to reach Bonferroni-corrected significance) for all 763,265 CpGs included in the analysis. CpGs were mapped to genes using an annotation object generated by the ‘IlluminaHumanMethylationEPICanno.ilm10b4.hg19’ R package (version 3.13)^22^. We searched the EWAS Atlas (https://ngdc.cncb.ac.cn/ewas/atlas) and EWAS catalog (http://www.ewascatalog.org/) for significantly associated CpGs and genes and the GWAS Catalog (https://www.ebi.ac.uk/gwas/) for annotated genes.

#### Pathway enrichment analysis

We used the ‘gometh’ function from the ‘missMethyl’ R package^23^ for pathway analyses using the results of the AHRR-adjusted model. Significant CpGs found in the MWAS after Bonferroni correction were selected, and the rest of the CpGs profiled in the EPIC array were included as the background list. Gene Ontology (GO) terms and Kyoto Encyclopedia of Genes and Genomes (KEGG) pathways were analysed separately. P-values for both enrichment analyses were FDR corrected.

#### Identification of differentially methylated regions (DMR)

DMR were analysed using the ‘dmrff’ R package^24^. A DMR was identified if contained at least two CpG sites within a maximum of 500-bp window, showed consistent direction of effect and both/all had meta-MWAS p < 0.05. Statistics were meta-analysed within the identified region. P-values were Bonferroni-corrected for all regions (>2 CpG sites) and single CpG sites altogether. Significant DMR were identified if the Bonferroni-corrected p < 0.05 and the number of CpG sites within the region was ≥ 2.

#### Analysis of the confounding effect of body mass index (BMI) and alcohol consumption

BMI and alcohol consumption are risk factors for MD and are known to have widespread associations with DNAm^7,25^. To investigate whether the signals found in the main model were due to the effects of BMI and alcohol consumption, we conducted an additional meta-analysis of a ‘complex model’ for the 14 cohorts that had BMI and alcohol consumption data available (N_total_ = 20,196, see Supplementary Figure 2 for sample sizes of individual studies). BMI and alcohol consumption were added as additional covariates for the complex model. Effect sizes and p values for the significant associations between the basic model and complex model were compared.

#### Out-of-sample classification of MD using MD-MS

##### Calculation of MD-MS and statistical model

We created MD-MS in an independent testing sample by calculating the weighted sum of M-values. Effect sizes from the MD meta-MWAS summary statistics were used as weights. Five p-value thresholds were used to select the CpG sites: p≤1, ≤0.01, ≤0.001, ≤1×10^-6^, and ≤5×10^-8^, resulting in five MD-MS.

MD diagnosis was set as the independent variable and MS as the dependent variable in the logistic regression model using the ‘glm’ function in R. Covariates were age, sex and M-values of AHRR probe. DNAm-estimated cell proportions were not associated with any of the MD-MS (p>0.5) and were therefore not included as covariates. Batch and genomic relationship matrix were pre-corrected by residualizing M-values against these covariates.

##### Testing sample

Generation Scotland (GS) DNAm data Set 3 was used for out-of-sample classification (GS DNAm data Set 1 and 2 were included in the meta-MWAS). GS DNAm data Set 3 used the Illumina Infinium Methylation EPIC array, had no overlap with the GS DNAm data used in the meta-MWAS, and relatedness within Set 3 and with the rest of the GS sample was removed by regressing the M-values against genomic relationship matrix. Quality check and preprocessing were kept consistent with the GS sample used in the meta-MWAS (Set 1 and 2).

MD diagnosis was derived using electronic health records (EHR) from General Practitioners’ (GP) diagnosis^26^. Details for the EHRs were explained in the protocol paper by Kerr *et al*^26^. In brief, a subset of participants (N = 20,032) of GS with genotyping data gave consent to link their data to EHRs. All Read codes from the PH1021 (https://phenotypes.healthdatagateway.org/phenotypes/PH1021/version/2204/detail/)^27,28^ and PH149 (https://phenotypes.healthdatagateway.org/phenotypes/PH149/version/298/detail/)^29^ inventories of the HDR UK Phenotype Library for primary care data of depression were used to identify cases of life-time MD. Participants with ≥ 1 entry of diagnosis of depression were classified as cases and those with no entry of any diagnosis for depression or no data to indicate depressive status were controls.

The final testing sample contained 504 cases and 8,372 controls.

##### Association between MD-MS and inflammatory protein markers

A previous study demonstrated wide-spread association between protein abundance and DNAm^30^. We conducted a proteome-wide association analysis study (PWAS) for MD-MS seeking to reveal the potentially downstream proteomic consequences of the measured DNAm differences.

Lothian Birth Cohort 1936 (LBC1936), a cohort independent of the MD MWAS, was used for the PWAS analysis. LBC1936 is a community-based birth cohort of participants born in 1936, recruited in Scotland. The sample used in the PWAS analysis had 875 people with both DNAm and proteome data collected at mean age 69.8±0.8_years. DNAm in LBC1936 was profiled in whole blood samples using the HumanMethylation450 BeadChip Kit (Ilumina, San Diego, CA, USA). Proteomic data was profiled using lithium heparin collected plasma samples analysed using a 92-plex proximity extension assay (inflammation panel), Olink® Bioscience, Uppsala Sweden. For 22 proteins over 40% of samples fell below the lowest limit of detection, leaving 70 post-quality-check proteins. Full details on sample preparation and quality check have been reported previously^31^.

General linear models (GLM) were used to test the association between relative abundance across all 70 proteins and MD-MS created at p-threshold ≤ 5×10^-8^. Protein abundance levels were residualised against age, sex, first four genetic PCs and array for proteomic data before entering association analysis. Residual scores of protein abundance were set as dependent variable. Array for DNAm data and the AHRR probe were included as covariates in the GLM. P-values were corrected using false discovery rate (FDR) correction.

#### Heterogeneity analysis

##### Leave-one-out analysis

To investigate whether a particular study had disproportionate influence on any meta-analytic association, we conducted leave-one-out meta-analyses. For each significant CpG, 18 iterations of meta-analysis were conducted leaving each individual study out, regardless of profiling arrays.

##### Meta-regression for age

For the CpGs that were significant in the MWAS meta-analysis, we used meta-regression to analyse the potential effect of age across studies. A mixed-effect model with the ‘metareg’ R package^32^ was used for the meta-regression analysis. Mean age of each individual cohort was set as a random effect. Standardised regression coefficients from summary statistics of each individual study were set as estimates of treatment effects (the ‘TE’ option). Between-study variance was estimated using the Restricted Maximum Likelihood (REML) method. The standardised regression coefficient of the random effect of age was reported as effect sizes for the meta-regression analysis.

##### Comparison between MD and BMI associations

To further evaluate the heterogeneity of MD-DNAm associations between studies, we looked at the correlation of effect sizes for the summary statistics of the MD MWAS for individual studies.

Generation Scotland (GS) that was included in the MWAS, being the largest study sample in the meta-analysis (N=9,502), was used to select a list of CpG sites of interest. The 1000 most significantly MD-associated CpG sites in GS were selected. Effect sizes for these CpG sites were extracted for all other studies. Correlation analysis was conducted on the effect sizes. We also performed a MWAS of BMI and conducted a similar analysis, for comparison with MD, to assess whether BMI-DNAm associations were similarly heterogeneous.

#### Mendelian randomisation (MR)

##### MD GWAS summary statistics

GWAS summary statistics were obtained from the Howard et al. meta-analysis for MD GWAS from the PGC, 23andMe and UK Biobank^3^. A total of 807,553 individuals (246,363 cases and 561,190 controls) of European ancestry were included in the MD meta-GWAS.

##### Genetics of DNA Methylation Consortium (GoDMC) and GS methylation quantitative trait loci (mQTL)

Quantitative trait loci associated with DNAm (mQTL) summary statistics were obtained from GS and GoDMC. For GS, full mQTL summary statistics (N=17,620) were obtained without any p-value thresholding. OmicS-data-based Complex trait Analysis (OSCA) was used for mQTL estimation^33^.

Details for the mQTL analysis in GS can be found elsewhere^34^. Covariates were kept consistent with the main model for the MD MWAS, except for using self-reported smoking behaviour (current smoker, past smoker, or non-smoker) and pack years (quantity of smoking) to control for smoking and adding ten genetic PCs as covariates. GoDMC mQTL data were obtained through the consortium website (http://www.godmc.org.uk/), using the same pipeline described in the GoDMC protocol paper by Min et al. (2021)^35^. The mQTL data contains 32 cohorts with 25,561 participants of European ancestry. The mQTL meta-analysis from GoDMC adopted a two-stage approach. First, a truncated set of mQTL data that reached the threshold of p-value < 1×10^-5^ were obtained from participating cohorts. This initial stage created a candidate list of mQTL associations (*n* = 120,212,413). Next, meta-analyses for mQTL were then conducted on these candidate associations. A full description of the GoDMC analysis can be found elsewhere^34,35^.

##### Samples for GS mQTL analysis, GoDMC mQTL analysis and MD GWAS were mutually exclusive

###### Selection of CpG list

A list of CpG sites that were either 1) significant in the MD MWAS or 2) within the identified DMRs were selected as CpG sites of interest. We extracted *cis* mQTL for further MR analysis^35^.

In the GoDMC dataset, a total of 156 CpG sites that met the above criteria had at least one *cis* mQTL. *cis* mQTL summary statistics for these CpG sites were extracted from the GoDMC dataset. For those CpG sites that had more than one *cis* mQTL after clumping (1 Mb window, p<5×10^-8^), the most significant mQTL with the lowest p-value was selected for analysis. Those CpG sites that showed significant causal association with MD were selected for replication analysis using the GS mQTL summary statistics.

###### MR methods

MR analysis was conducted using the ‘TwoSampleMR’ R package (version 0.5.6)^36^. To identify causal effects of DNAm on MD, we used the Wald’s ratio MR method^37^ to analyse causal effects on MD using cis mQTLs (within a 1Mb window in vicinity of the chosen CpG site). The most significant mQTL for each CpG site that reached the threshold of p<5×10^-8^ was selected. Causal effects from DNAm to MD were tested using both GoDMC and GS mQTL data. For the causal effect in the reverse direction (from MD to DNAm), MD GWAS summary statistics were clumped at p ≤ 5×10^-8^, with a 1Mb window and r=0.001. Causal effects from MD to DNAm were tested using mQTL data from the entire GS sample (Set 1, 2 and 3).

##### MWAS in East Asian ancestry

We sought to investigate MD associations with DNAm in participants of East Asian (Taiwan Biobank) ancestry. Demographic and descriptive statistics are included in the Supplementary Materials. As in the main meta-analysis, biological and technical covariates, as well as age, sex, and smoking (indexed by AHRR probe cg05575921) were included as covariates. Evidence of trans-ancestry transferability of MD-CpG effects was investigated by testing for the correlation of effect sizes in both ancestries. We then used the function “gometh” in package ‘missMethyl’ to assess ontology and pathway enrichment (GO and KEGG) for differentially methylated CpG sites at a threshold of_*p* < 1×10^−5^_(N_CpG_=24), as used in previous studies^38^.

## Results

### MD meta-MWAS

A total of 15 CpG sites were significant after Bonferroni correction from the basic model (p < 6.55×10^-8^, p_Bonferroni_ < 0.05, see Table 1 for details of participating studies, Figure 2 and Table 2 for significant findings).

**Table 2.**
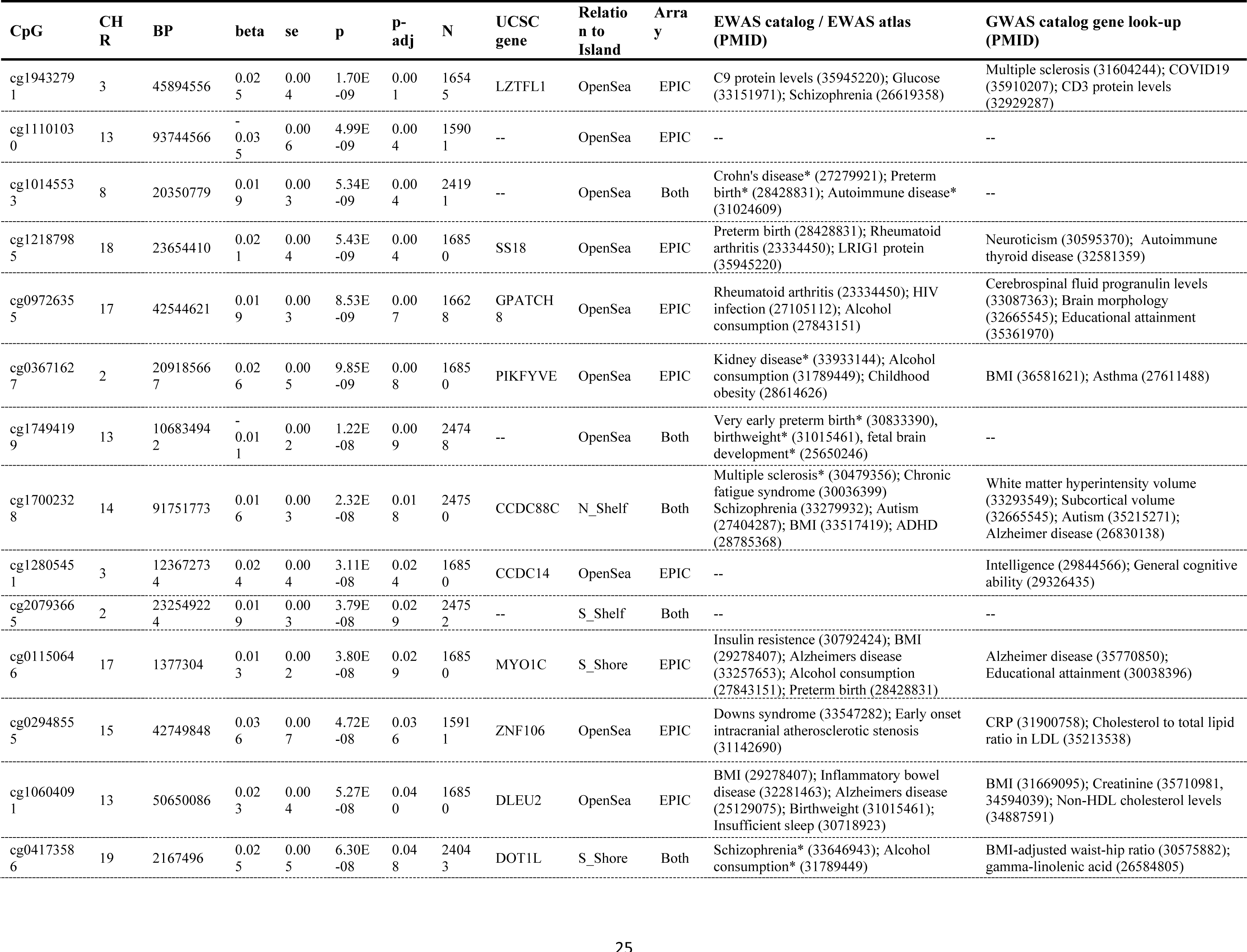
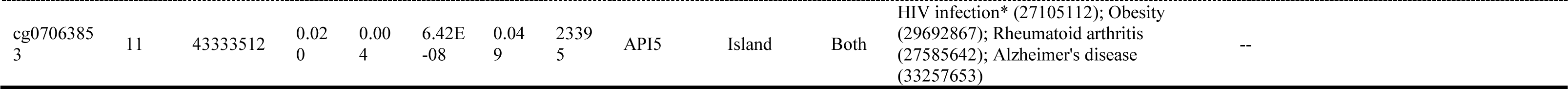
Significant CpG sites in the meta-analysis of MD MWAS. EWAS catalog (http://www.ewascatalog.org/), EWAS atlas (https://ngdc.cncb.ac.cn/ewas/atlas) and GWAS catalog (https://www.ebi.ac.uk/gwas/) were used to find associated traits in previous studies (traits associated with CpG sites are marked with *).

**Figure 2.**
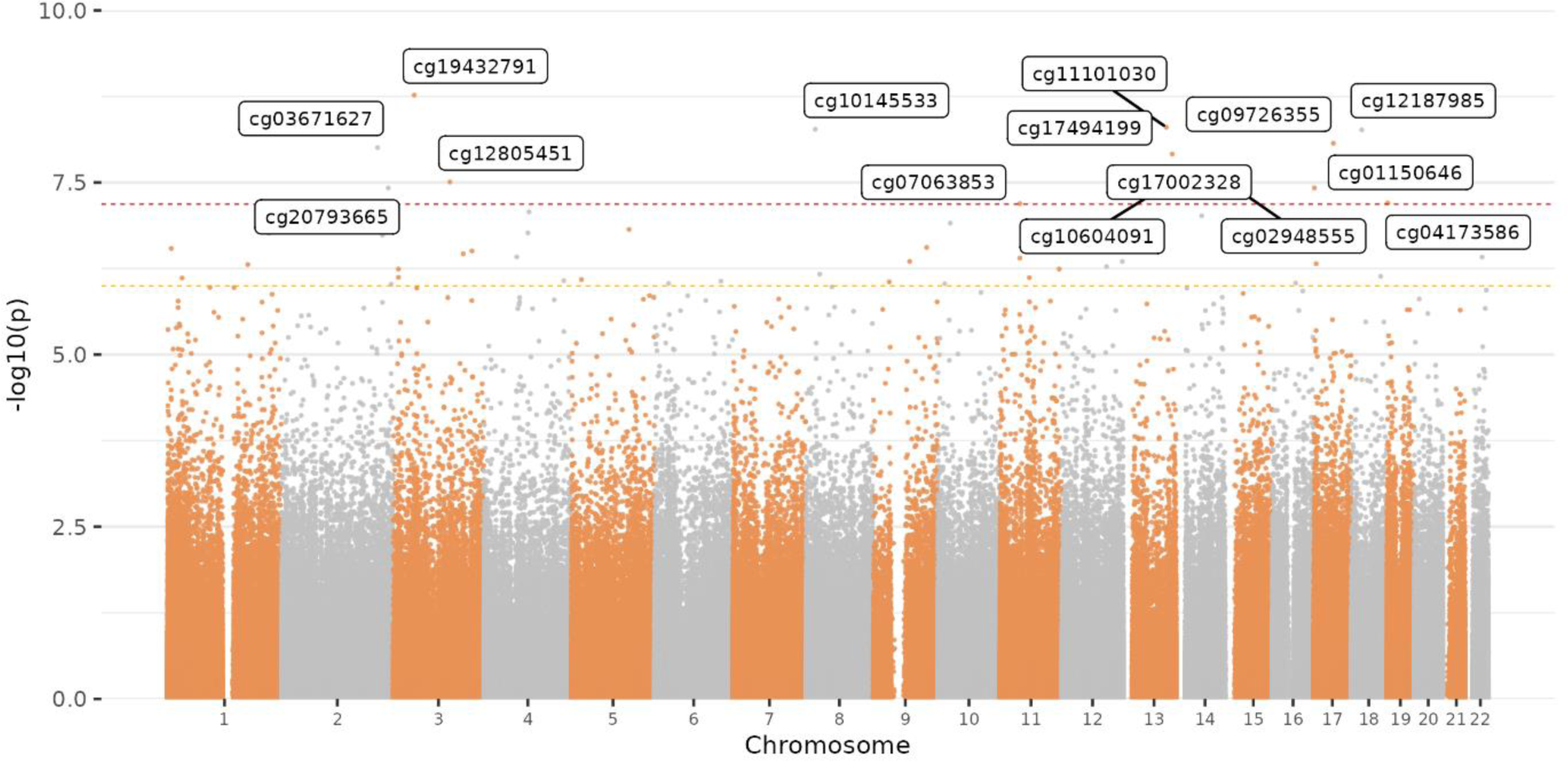
Manhattan plot for the meta-analysis of MWAS for MD. Each dot represents a CpG probe. X-axis represents the relative position of the probes in the genome. Y-axis represents −log10-transformed p-values. The red and yellow dashed line represents the significance threshold for Bonferroni and FDR correction, respectively.

Five of the significant CpG sites positionally map to genes associated with mental health, neurodegenerative and developmental disorders. The gene mapped from cg17002328 (*CCDC88C*) was associated with schizophrenia^39^ and ADHD^40^ in epigenetic studies, and brain structural measures in GWAS studies, such as cortical surface area^41^ and accumbens volume^42^. CpG site cg17494199 was associated with preterm birth^43^, birthweight^44^ and foetal brain development^45^ in previous epigenetic studies. Genes mapped from cg01150646 (*MYO1C*), cg10604091 (*DLEU2*) and cg07063853 (*API5*) were associated with Alzheimer’s disease in brain-tissue methylation levels^46–48^.

A total of five significant CpG sites were associated with autoimmune diseases and biomarkers in previous studies, one of which was the most significantly CpG site (cg19432791 on chromosome 3), mapping to gene *LZTFL1*. From the EWAS catalog, the *LZTFL1* gene was associated with biomarkers related to pain, such as glucose level^49^, and autoimmune disease or markers, such as rheumatoid arthritis^50^ and C9 protein levels^30^. Four of the significant CpG sites, cg12187985 (*SS18*), cg10145533, cg09726355 (*GPATCH8*) and cg07063853 (*API5*), map to genes previously found associated with autoimmune markers (e.g. rheumatoid arthritis^50^) in MWAS studies.

Other CpG sites, cg03671627 and cg02948555 (*ZNF106*), were associated with traits and markers relevant to obesity in both genetic and epigenetic studies. These markers include, for example, BMI^25^ and cholesterol to total lipids ratio^51^.

A complete list of CpG sites and related genes and traits can be found in Table 2. The inflation factors of the association statistics for each individual cohort can be found in Supplementary Figure 3. See Supplementary Figure 4 for effect sizes of each study for the significant CpG sites.

### Identification of DMR

A total of 37 DMRs were identified after Bonferroni correction. The most strongly associated region locates within the major histocompatibility complex (MHC) region, discoidin domain receptor tyrosine kinase 1 (*DDR1)* gene (CHR6:30,853,258-30,854,233), previously implicated in major psychiatric disorders (e.g. schizophrenia, MD and bipolar disorder).

The top ten most significant DMRs are listed in Table 3, and the full list of significant DMRs can be found in Supplementary Table 1.

**Table 3.** Top ten significant differentially methylated regions (DMRs) identified in the basic model. Start/End = start/end base pair position of the DMR.

| CHR | Start | End | Gene(s) | N<sub>CpG</sub> | Beta | se | p | p.bonferroni |
| --- | --- | --- | --- | --- | --- | --- | --- | --- |
| 6 | 30853258 | 30854233 | DDR1 | 12 | 0.036 | 0.005 | 9.89E-13 | 6.75E-08 |
| 19 | 37825211 | 37825455 | HKR1 | 8 | 0.036 | 0.005 | 5.66E-12 | 3.87E-07 |
| 10 | 135342560 | 135343248 | CYP2E1 | 5 | 0.031 | 0.005 | 1.42E-10 | 9.68E-06 |
| 3 | 18480242 | 18481064 | SATB1 | 8 | 0.048 | 0.009 | 1.16E-07 | 0.008 |
| 6 | 30698784 | 30698936 | FLOT1 | 4 | 0.046 | 0.009 | 1.45E-07 | 0.010 |
| 11 | 43333495 | 43333512 | API5 | 3 | 0.055 | 0.009 | 2.59E-09 | 1.77E-04 |
| 17 | 7832769 | 7832932 | KCNAB3 | 3 | 0.036 | 0.005 | 1.14E-13 | 7.78E-09 |
| 9 | 139557250 | 139557920 | EGFL7 | 4 | -0.059 | 0.011 | 2.20E-08 | 0.002 |
| 2 | 219738714 | 219738732 | WNT6 | 2 | 0.074 | 0.014 | 5.44E-08 | 0.004 |
| 3 | 11643427 | 11643630 | VGLL4 | 3 | 0.083 | 0.015 | 4.45E-08 | 0.003 |

### Pathway enrichment analysis

No GO term or KEGG pathway was significantly enriched after FDR correction (Supplementary Table 2). The top 10 most significantly enriched GO terms included pathways relevant to protein and metabolic processes (e.g. negative regulation of protein localisation to ciliary membrane) (p ranged from 0.003 to 8.14×10^-4^). For KEGG, ‘transcriptional misregulation in cancer’ and ‘lysine degradation’ were the only pathways that reached nominal significance (p ≤ 0.007).

### The basic model versus complex model

Results for the basic model (18 cohorts, N_total_ = 24,754) and complex model (15 cohorts, N_total_ = 20,196, see Supplementary Figure 2) were compared to evaluate the confounding effect of BMI and alcohol consumption (Figure 3). Effect sizes of the two models were highly correlated for the significant CpG sites (r=0.988) and for CpG sites across the entire methylome (r = 0.920). All significant CpG sites in the basic model remained significant in the fully adjusted model after Bonferroni correction across the 15 significant CpGs found in the basic model, despite a significant reduction in sample size (p < 2.51×10^-4^, p_Bonferroni_ < 0.004 corrected across the significant CpG sites in discovery analysis).

**Figure 3.**
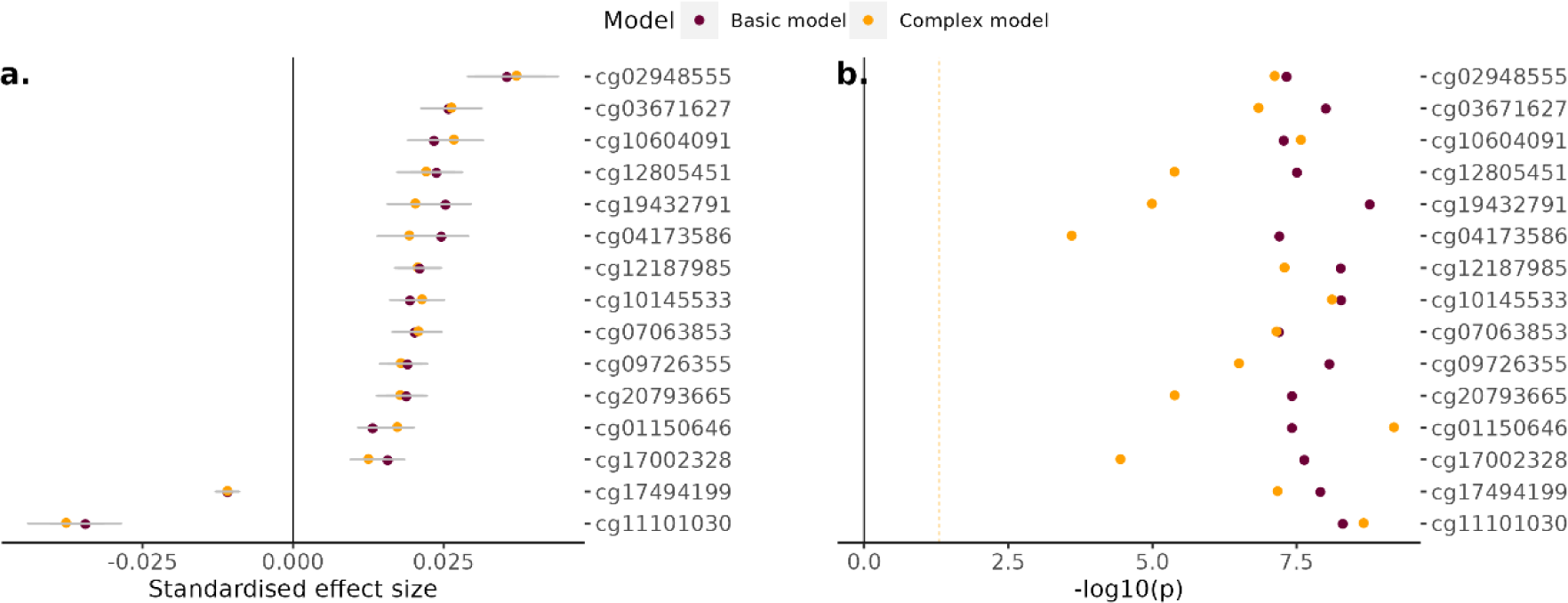
Comparison between basic and complex models. In both (a) comparison of effect sizes and (b) comparison of p values, each dot represents a CpG probe. In panel (a), x-axis represents effect size, and in panel (b), x-axis represents −log10-transformed p value. Differences in p-values reflect both the model used and the sample sizes. Y-axis represents individual CpG site. The yellow dashed line in panel (b) represents the significance threshold for nominal significance (p < 0.05).

### Out-of-sample classification of MD using MD-MS

All five MD-MS showed positive effect sizes (higher score associated with higher liability of MD). There was an increasing trend of effect sizes as the p-value threshold for MS calculation becomes increasingly stringent (see Figure 4). Out of the five scores tested, only MS at p-value threshold of ≤5×10^-8^ was found associated with MD diagnosis (β = 0.13, p = 0.003, see Figure 4). P-values for other MS ranged from 0.069 to 0.314.

**Figure 4.**
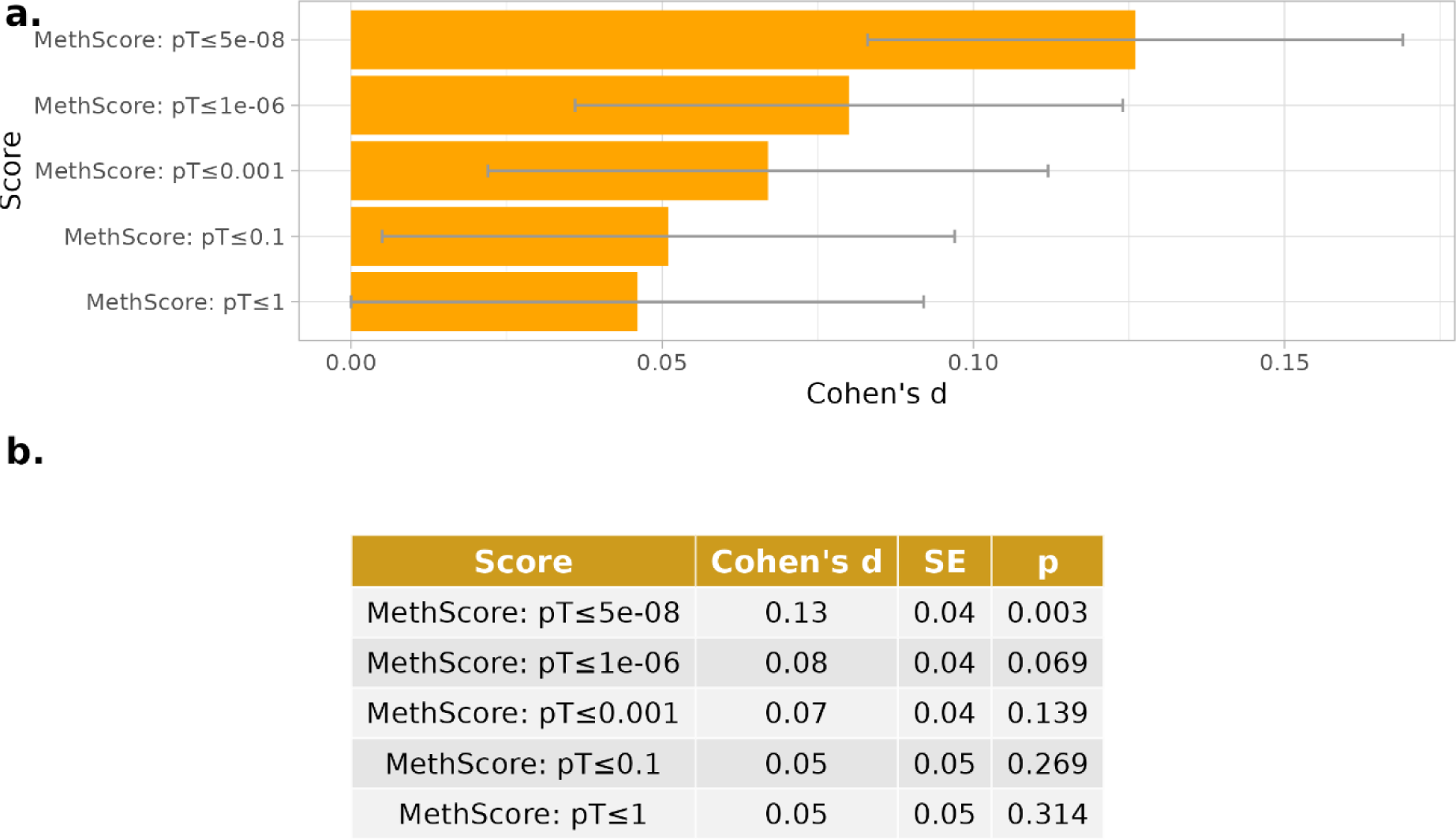
Out-of-sample classification of MD using MS. (a) Bar-plot for effect sizes of associations. Length of bar represents standardised log-transformed odds ratio and standard error (error bar). Y-axis represents MS. (b) Statistics for MS association tests.

### Association between MS for MD and inflammatory protein abundance

Five proteins were significantly (p < 3.5×10^-3^) associated with the MD-MS created at the p-threshold of ≤5×10^-8^. The strongest association was found in *TNFB* (β = −0.15, p = 2.37×10^-5^). Other proteins were: *IL6* (β = 0.12, p = 5.18×10^-4^), *TFG alpha* (β = 0.11, p = 2.8×10^-3^), CD5 (β = −0.11, p = 2.1×10^-3^) and *EN-RAGE* (β = 0.11, p = 3.5×10^-3^). See Supplementary Figure 5.

### Heterogeneity analysis

#### Leave-one-out analysis

For every CpG site, all effect sizes for leave-one-out analyses remained in the same direction as the meta-analysis (Supplementary Figure 6). All tested CpG sites remained significant after leaving individual studies out except when the largest study, Generation Scotland, was left out (p < 1.03×10^-4^). When Generation Scotland was left out, 11 out of the 15 tested CpG sites remained nominally significant (p < 8.91×10^-3^). The four sites that became non-significant when Generation Scotland was omitted were: cg02948555 (β_meta-MWAS_ = 0.036, β_LOO_ = 0.008, p_LOO_ = 0.511), cg11101030 (β_meta-MWAS_ = −0.035, β_LOO_ = −0.01, p_LOO_ = 0.235), cg07063853 (β_meta-MWAS_ = 0.02, β_LOO_ = 0.01, p_LOO_ = 0.232) and cg01150646 (β_meta-MWAS_ = 0.013, β_LOO_ = 0.007, p_LOO_ = 0.054).

#### Effect of age difference between studies

Out of the 15 CpG sites significant in the meta-MWAS, 14 sites did not show an effect of age difference across studies (absolute β ranged from 1.11×10^-6^ to 6.09×10^-4^, p >0.234, Supplementary Figure 7). One site, cg04173586, showed a significant effect of age (β = −9.25×10^-4^, p =0.006).

However, leave-one-out analysis both showed highly consistent findings for cg04173586 across studies. There were 11 out of 15 participating cohorts that showed effect sizes consistent with the meta-MWAS (Supplementary Figure 7) and leave-one-out analyses were significant for all iterations (p ranged from 1.03×10^-4^ to 2.38×10^-8^, Supplementary Figure 4 and Supplementary Table 3).

#### Correlation matrix for effect sizes

Heterogeneity between studies was analysed by looking at the between-study correlation of effect sizes estimated using the basic model (see Figure 5). Correlations between the 18 studies participating in the MD meta-MWAS ranged from −0.19 to 0.31. The highest positive correlation was found between the Netherland Twin Register and Janssen (r=0.31). Out of the 153 pairwise correlations, 96 were positive (62.7%). Compared to the MD meta-MWAS, BMI MWAS (10 studies) showed higher, positive effect size correlations between studies (r ranged from 0.305 to 0.864, 100% of the pairs were positively and significantly correlated).

**Figure 5.**
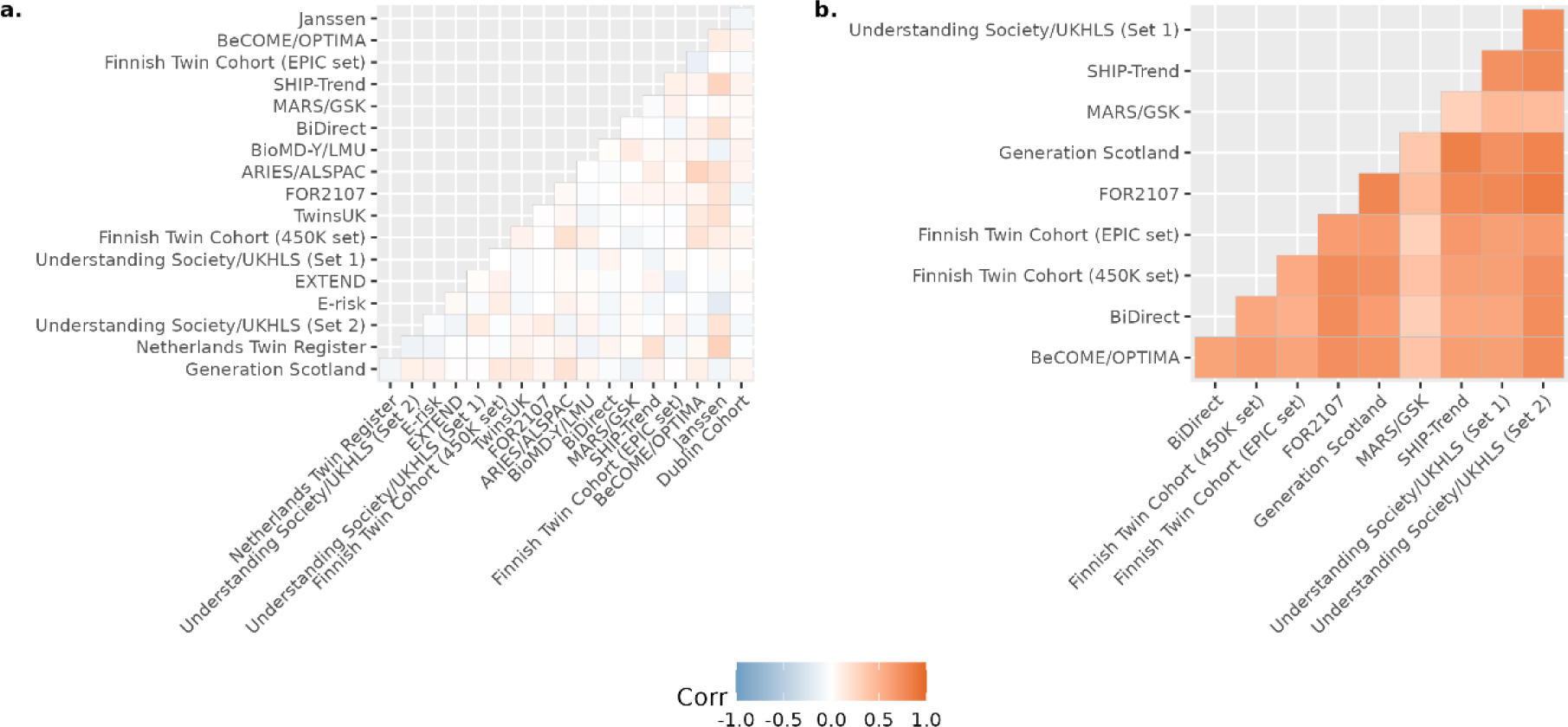
Correlation matrices for MD and BMI MWAS, respectively. Panel (a) shows the correlation matrix for the top 1000 CpG sites in GS MD MWAS. Pair-wise correlation of effect sizes are shown in the figure. Panel (b) shows the correlation matrix for the top 1000 CpG sites in GS BMI MWAS. Pair-wise correlation of effect sizes are shown in the figure.

### Mendelian Randomisation (MR)

Using *cis* mQTL data from GS and Wald’s ratio MR method, we found 23 significant and potentially causal effects of DNA methylation on MD (absolute β ranged from 0.06 to 0.93, p ranged from 6.88×10^-3^ to 4.58×10^-6^, see Figure 6a and Supplementary Data 1). There were 17 CpG sites located in the MHC region (mapping to the *DDR1R* gene) on liability to MD (β ranged from 0.06 to 0.17, p ranged from 1.1×10^-3^ to 8.05×10^-5^). See Supplementary Data 1 for the full list of significant causal effects found in the discovery analysis in GS.

**Figure 6.**
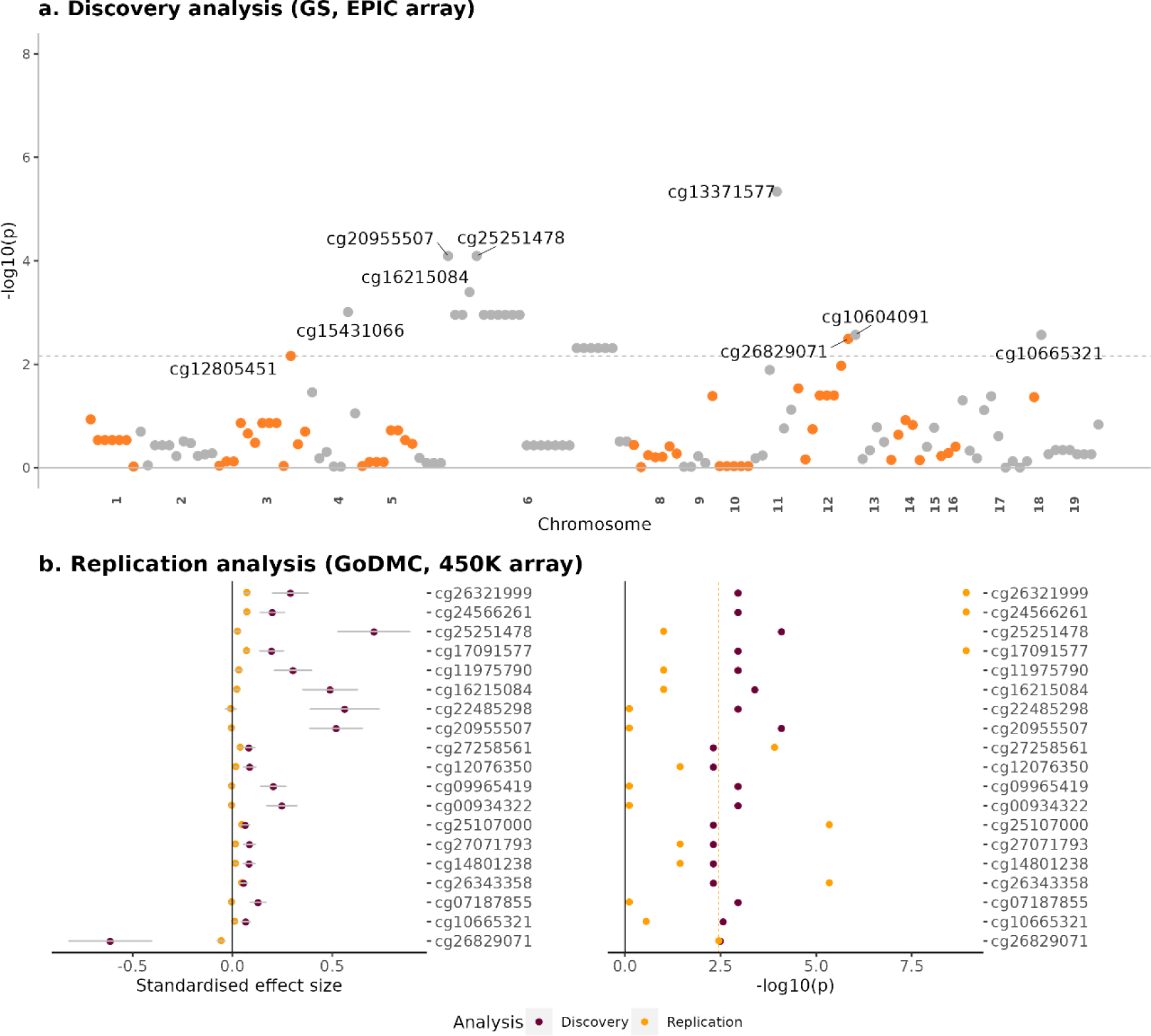
Significance of Mendelian Randomisation analysis of the causal effect of DNAm on MD. (a) P-plot for discovery MR analysis in GS. Each dot represents a CpG site. X-axis represents chromosomes and base pair position. Y-axis represent −log10-tranformed p-value of Wald’s ratio MR analysis. The grey dashed line shows the FDR-corrected significance threshold. Discovery MR analysis was performed on CpG sites available in the EPIC array. (b) Replication analysis in GoDMC for the significant CpGs found in the discovery analysis. Replication analysis was performed on CpG sites available on the 450K array. Out of the 23 significant CpG sites in the discovery analysis, 19 were available on both EPIC and 450K arrays and therefore were included in the replication analysis. X-axes represent effect size and −log10-transformed p value on the left and right panels. Y-axis represents individual CpG site. The yellow, dashed line in the right panel represents FDR-significance threshold.

Out of the 23 potentially causal effects found in the discovery analysis, four CpG sites were specific to the EPIC array, while 19 were available on both array types and could, therefore, be included in the replication analysis. Seven CpG sites tested were replicated (p<0.003, Figure 6b). Six of the replicated CpG sites were in the MHC region (β: 0.047 to 0.073, p: 1.22×10^-4^ to 1.19×10^-9^). In addition, a potentially causal effect from cg26829071 to MD (mapping to *GPR133*, β = −0.056, p = 0.004) was found on chromosome 12. No significant causal effects from MD to DNAm were found (absolute β: 1.62×10^-5^ to 0.22, p_uncorrected_ > 0.001, p_FDR_ > 0.4).

### MWAS in East Asian ancestry

We sought to identify DNAm associations with MD in East Asian ancestry using data from Taiwan Biobank (TBB). There were no methylome-wide significant findings identified. Correlation between the methylome-wide significant CpGs identified in the main results (N=15) and the same CpGs in TBB was r = 0.48. Effect direction across the two cohorts was the same for 11 of the 15 CpGs (73.3% in the same direction, see Supplementary Table 4-6). Positive correlations of effect sizes when we extended the comparison in the top 100 and top 1,000 CpG sites found within the MWAS from European samples (top 100 CpG sites found in European samples: r = 0.26, p = 0.009; top 1,000 CpG sites found in European samples: r = 0.22, p = 4.07×10^-12^).

## Discussion

Our meta-MWAS of 24,754 individuals found 15 CpG sites to be significantly associated with MD, an increase of 12 CpGs on previously reported associations^11^. Significant findings revealed CpGs mapped to genes associated with autoimmune markers and depression-related traits, such as BMI. Seven potentially causal effects from DNAm to MD were identified and replicated. Leave-one-out analysis showed that effects were highly consistent across studies for the significant sites. However, the correlation in CpG-MD association effect sizes was relatively heterogeneous and low for MD, in contrast to BMI which showed uniformly positive, significant and more homogeneous correlations across all available studies. A positive correlation was observed for top MD CpG sites between European and East Asian samples (r = 0.48 for significant CpG sites and r = 0.26 for top 100 CpG sites), and effects of 11 of the 15 significant CpG sites found in the European sample remained in the same direction in the East Asian sample.

Five CpG sites mapped to genes previously reported in association with autoimmune disorders and markers relevant to activity of the hypothalamic-pituitary-adrenal (HPA) axis. The top CpG, cg19432791, mapped to *LZTFL1*, a gene that was identified as an effector gene that contributes to severe autoimmune responses and inflammation, such as risk of respiratory failure caused by COVID-19^52^. In addition to its association with inflammation, *LZTFL1* regulates ciliary localisation of the Bardet–Biedl syndrome (BBSome) protein complex. The BBSome complex is a well-replicated causative protein marker for obesity^53^ and highly relevant to the HPA-axis activity by being involved in transducing leptin signals in hypothalamic neurons^54,55^. Consistent with the top CpG site, other sites, such as cg02948555 and cg10604091, mapped to genes associated with C-reactive protein (CRP)^8,56^. CRP is one of the most well-studied predictors of vulnerability to depression^57^ and persistence of depressive symptoms^58^. As a pro-inflammatory cytokine, CRP has been found as an indicator of hyper-activity in the HPA-axis associated with stress^59^. The PWAS findings provided additional evidence for the association between the top CpG sites and inflammation, for example, in tumor necrosis factor-beta (*TNFB*) and Interleukin 6 (*IL6*). Our findings, provide further evidence supporting the association between MD and chronic inflammation, particularly involving the HPA-axis. However, no association between these results and MD had been identified in previous MWAS, likely due to the limited sample sizes of MWAS conducted on mental health-related traits.

Three CpG sites were associated with traits (e.g blood cholesterol levels, waist-to-hip ratio and pulse pressure) related to BMI. Obesity has been repeatedly associated with depression, and MR analyses from previous studies have indicated that this may be a causal association^3,60^. Within the genes in vicinity of the four CpG sites, *ZNF106* is involved in the regulation of calcium homeostasis that is crucial for cell survival and death^61^. Other genes, including *MYO1C* and *FNDC3B*, are understood to be involved in energy metabolism and homeostasis and highly sensitive to stress coping and inflammation^62,63^. Although the previous findings relevant to the BMI-related genes were based on GWAS studies, one of the genes, *FNDC3B*, was found in an earlier MD MWAS study based on brain tissue samples^13^. Taking into account the dynamic nature of DNA methylation and its high sensitivity to environmental stressors, our findings suggest metabolic processes may play a potentially crucial role in depression, which may be exacerbated by adverse environmental factors and dysfunctional stress coping mechanisms.

We observed varying results from different cohorts in our study, with little evidence of systematic differences in age differences between studies contributing to the heterogeneity. The higher degree of effect size heterogeneity in the MD versus BMI MWAS meta-analyses suggests that phenotype may be a reason for differences between the studies and not other methodological factors, such as sample processing or covariate adjustment. In the MD meta-analysis, larger studies (N > 1000) showed stronger correlation for the top associations, suggesting that sufficient statistical power may help overcome the issue of phenotyping inconsistency. This suggests that future individual MWAS studies of MD should be larger in size. The considerable costs of DNAm profiling, compared to genotyping, unfortunately act as a barrier towards achieving these aims.

Although there was no significant finding in the MWAS conducted in non-European studies, it is notable that effects were consistent with the significant findings on European samples. Top pathways associated with MD also showed convergent results with the main analysis (e.g. pathways relevant to inflammation and immune processing). Correlations for effect sizes between European and East Asian samples were positive with the South Asian sample, consistent with previous evidence showing relatively high genetic consistency between European and East Asian groups^64^. Developing larger non-European DNA methylation samples will be crucial to give statistical balanced comparison between ancestry groups and to identify ancestry-group-specific DNA methylation sites for MD^65^.

Our study combined many studies with widely varying sample sizes, compared results between two ancestry groups, and replicated MR findings using two large mQTL datasets. We provided a comprehensive evaluation of sampling and analytic strategies to guide future large-scale meta-MWAS for mental health disorders. While the blood draw was not timed to coincide with the onset of a depressive episode, limiting causal inferences based on the temporal order of DNAm exposure and MD onset, the MR analyses helped to address this potential limitation. Additionally, there is the lack of replication in other tissue types that are directly relevant to mood regulation, such as brain tissue. Studies have shown that the genetic drivers of DNAm have similar effects across multiple cell types^66^. Future clinical applications and larger sample sizes make whole-blood DNAm data more feasible than post-mortem tissue samples. However, to ensure the validity of the findings, future studies should broaden their scope by encompassing additional cell and tissue types.

## Funding and acknowledgement

### Generation Scotland (GS)

This work is supported by three Wellcome Trust grants (220857/Z/20/Z, 104036/Z/14/Z, 216767/Z/19/Z). DNAm profiling was supported by funding from NARSAD (Ref 27404, DMH) and the Royal College of Physicians of Edinburgh (SIM Fellowship, HCW). Genotyping of the GS samples was funded by the MRC and Wellcome Trust [104036/Z/14/Z]. GS also receives support from the Chief Scientist Office of the Scottish Government Health Directorates [CZD/16/6] and the Scottish Funding Council [HR03006].

### Avon Longitudinal Study of Parents and Children (ALSPAC) / Accessible Resource for Integrated Epigenomic Studies (ARIES)

The UK Medical Research Council and Wellcome (Grant Ref: 217065/Z/19/Z) and the University of Bristol provide core support for ALSPAC. A comprehensive list of grants funding is available on the ALSPAC website (http://www.bristol.ac.uk/alspac/external/documents/grant-acknowledgements.pdf). Part of this data was collected using REDCap, see the REDCap website for details https://projectredcap.org/resources/citations/). We thank all the families who took part in this study, the midwives for their help in recruiting them, and the whole ALSPAC team, which includes interviewers, computer and laboratory technicians, clerical workers, research scientists, volunteers, managers, receptionists and nurses.

### BeCOME/OPTIMA

#### Funding

OPTIMA and BeCOME are funded by the Max Planck Society.

#### Acknowledgements

We would like to thank all contributors to the research project including physicians, psychologists, study nurses, researchers and research assistants, and of course patients of the hospital of the Max Planck Institute of Psychiatry in Munich.

### BiDirect

The BiDirect Study is supported by grants of the German Ministry of Research and Education (BMBF) to the University of Münster (01ER0816 and 01ER1506).

### Biopsychosocial factors of major depression in youth (BioMD-Y/LMU)

BioMD-Y was supported by the Randebrock Stiftung associated with the kbo-Heckscher-Klinikum, Munich, Germany during the years 2009-2011 upon its inception, and has since been financed through intramural funds. DNA extraction, genome-wide genotyping and DNA methylation analyses were supported by the Max Planck Society. DNA Methylation of the BioMD-Y/LMU cohort was funded in part by Sabine Schaefer, private Donor of the Max-Planck-Foundation.

### Dublin cohort

This research was supported by a grant from AbbVie (no. 10118) to HM, TMM and EMcD. EMcD was the recipient of the Boston Scientific Newman Fellowship awarded by the UCD Foundation. The authors wish to thank all the participants in the Dublin Cohort and all researchers involved in the original IBD Study.

### Environmental Risk (E-Risk) Longitudinal Twin Study

The E-Risk Study is funded by grants from the UK Medical Research Council [G1002190; MR/X010791/1]. Additional support was provided by the US National Institute of Child Health and Human Development [HD077482] and the Jacobs Foundation. This project represents independent research part funded by the NIHR Maudsley Biomedical Research Centre at South London and Maudsley NHS Foundation Trust and King’s College London. The views expressed are those of the authors and not necessarily those of the NIHR or the Department of Health and Social Care [NIHR203318]. Helen L. Fisher was supported by the UK ESRC Centre for Society and Mental Health at King’s College London [ES/S012567/1]. The views expressed are those of the authors and not necessarily those of the ESRC or King’s College London.

We are grateful to the E-Risk study mothers and fathers, the twins, and the twins’ teachers for their participation. Our thanks to the E-Risk team for their dedication, hard work, and insights.

### EXTEND

This study was funded by the National Institute for Health and Care Research Exeter Clinical Research Facility. This study was supported* by the National Institute for Health and Care Research Exeter Biomedical Research Centre. The views expressed are those of the author(s) and not necessarily those of the NIHR or the Department of Health and Social Care.

### Finnish Twin Cohort

Phenotype and genotype data collection in FinnTwin12 and FinnTwin16 studies of the Finnish twin cohort has been supported by the Wellcome Trust Sanger Institute, the Broad Institute, ENGAGE – European Network for Genetic and Genomic Epidemiology, FP7-HEALTH-F4-2007, grant agreement number 201413, National Institute of Alcohol Abuse and Alcoholism (grants AA-12502, AA-00145, and AA-09203 to R J Rose; AA15416 and K02AA018755 to D M Dick; R01AA015416 to Jessica Salvatore) and the Academy of Finland (grants 100499, 205585, 118555, 141054, 264146, 308248 to JK, 328685, 307339, 297908 and 251316 to MO, 309119 to TK) and Sigrid Juselius Foundation to MO and JK. and the Centre of Excellence in Complex Disease Genetics (grants 312073, 336823, and 352792 to JKaprio). JKaprio acknowledges support by the Academy of Finland (grants 265240, 263278). Tellervo Korhonen acknowledges support from the Juho Vainio Foundation.

We would like to thank the active participation of the twins, which has made the studies possible.

### FOR2107

The German multicenter consortium “Neurobiology of Affective Disorders. A translational perspective on brain structure and function” is funded by the German Research Foundation (Research Unit FOR2107). Principal investigators are Tilo Kircher (KI588/14-1, KI588/14-2,, KI588/20-1, KI588/22-1), Udo Dannlowski (DA1151/5-1, DA1151/5-2), Axel Krug (KR3822/5-1, KR3822/7-2), Igor Nenadic (NE2254/1-2,NE2254/3-1,NE2254/4-1), Carsten Konrad (KO4291/3-1), Marcella Rietschel (RI 908/11-1, RI 908/11-2), Markus Nöthen (NO 246/10-1, NO 246/10-2), Stephanie Witt (WI 3439/3-1, WI 3439/3-2). Tilo Kircher received unrestricted educational grants from Servier, Janssen, Recordati, Aristo, Otsuka, neuraxpharm. We are deeply indebted to all study participants and staff. A list of acknowledgments can be found here: www.for2107.de/acknowledgements.

### Janssen

#### Funding

The Janssen cohort was funded by Janssen Research & Development, LLC.

#### Acknowledgements

We thank the clinical investigators and research coordinators who ran the clinical study and collected the blood samples used in this study, as well as study participants and their families, whose help and participation made this work possible. We thank the staff from Cancer Genetic Institute and HD Bioscience for performing the DNA extraction from whole blood samples, plating, QC, and the staff from Illumina to perform the epigenetic and genotyping assays to enable the data generation.

### Munich Antidepressant Response Signature (MARS/GSK)

#### Funding

The MARS cohort was sponsored by the Max Planck Society. The UniDep cohort was funded by the Bavarian Ministry of Commerce and by the Federal Ministry of Education and Research in the framework of the National Genome Research Network, Foerderkennzeichen 01GS0481 and the Bavarian Ministry of Commerce. DNA methylation analysis of a subset of both cohorts was financed by ERA-NET NEURON.

#### Acknowledgements

We would like to thank all contributors to the research project including physicians, psychologists, study nurses, researchers and research assistants, and of course patients of the hospital of the Max Planck Institute of Psychiatry in Munich and psychiatric hospitals in Augsburg and Ingolstadt.

### Lothian Birth Cohort 1936 (LBC1936)

The LBC1936 is supported by the Biotechnology and Biological Sciences Research Council, and the Economic and Social Research Council [BB/W008793/1] (which supports SEH), Age UK (Disconnected Mind project), the Medical Research Council [G0701120, G1001245, MR/M013111/1, MR/R024065/1], the Milton Damerel Trust, and the University of Edinburgh. SRC is supported by a Sir Henry Dale Fellowship jointly funded by the Wellcome Trust and the Royal Society (221890/Z/20/Z). Methylation typing of was supported by Centre for Cognitive Ageing and Cognitive Epidemiology (Pilot Fund award), Age UK, The Wellcome Trust Institutional Strategic Support Fund, The University of Edinburgh, and The University of Queensland.

### Netherlands Twin Register (NTR)

We warmly thank all twin families of the Netherlands Twin Register who make this research possible. This work was supported by the Royal Dutch Academy for Arts and Science (KNAW) Academy Professor Award (PAH/6635) to DIB; the Netherlands Organization for Scientific Research (NWO 480-15-001/674) and Biobanking and Biomolecular Resources Research Infrastructure (BBMRI-NL: 184.021.007; 184.033.111).

### Study of Health in Pomerania (SHIP)

SHIP is part of the Community Medicine Research net of the University of Greifswald which is funded by the Federal Ministry of Education and Research (01ZZ9603, 01ZZ0103, and 01ZZ0403), the Ministry of Cultural Affairs and the Social Ministry of the Federal State of Mecklenburg-West Pomerania. DNA methylation data have been supported by the DZHK (grant 81X3400104). The University of Greifswald is a member of the Caché Campus program of the InterSystems GmbH. The SHIP authors are grateful to Paul S. DeVries for his support with the EWAS pipeline.

### Taiwan Biobank

This work is supported by Population Health Research Center from Featured Areas Research Center Program within the framework of the Higher Education Sprout Project by the Ministry of Education in Taiwan (grant number NTU-112L9004).

We thank all the participants and investigators of the Taiwan Biobank.

### TwinsUK

The study received support from the ESRC (ES/N000404/1 to J.T.B) and European HDHL Joint Programming Initiative funding scheme DIMENSION project (BBSRC BB/S020845/1 and BB/T019980/1 to J.T.B.). The TwinsUK study is funded by the Wellcome Trust, Medical Research Council, Versus Arthritis, European Union Horizon 2020, Chronic Disease Research Foundation (CDRF), ZOE LIMITED, and the National Institute for Health Research (NIHR) Clinical Research Network (CRN) and Biomedical Research Centre based at Guy’s and St Thomas’ NHS Foundation Trust in partnership with King’s College London.

### Understanding Society/ UK Household Longitudinal Study (UKHLS)

We acknowledge the Wellcome Trust Sanger Institute and Ele Zeggini for gen
erating the genotype data and the School of Life Sciences, University of Essex for QC of the methylation data. Both genotyping and DNA methylation in UKHLS were funded through enhancements to the Economic and Social Research Council (ESRC) grants ES/K005146/1 and ES/N00812X/1. MK is partially supported by the ESRC (ES/S012486/1). AD and ERW are Soc-B students (ESRC project reference numbers 2765580 and 2604212 respectively).

We would also like to acknowledge the summary statistics provided by the Psychiatric Genomics Consortium. In addition, we would like to thank the research participants and employees of 23andMe, Inc. for making this work possible.

## Data Availability

All summary statistics produced in the present study are available upon reasonable request to the authors upon publication. Individual-level data requires approval of proposed analyses and data transfer agreement (if applicable).

