## Supplementary Materials for "A methylome-wide association study of major depression with out-of-sample case-control classification and trans-ancestry comparison"

**MWAS for major depression**

**Data preparation for each individual study**

**Generation Scotland (GS)^1^**

**DOI for Protocol paper:** <https://doi.org/10.12688/wellcomeopenres.15136.1>

MD phenotype: Baseline MDD status was measured using the axis-I Structured Clinical Interview of the Diagnostic and Statistical Manual, version IV (SCID) and was administered to participants who answered “yes” to either of two screening questions. MDD status was measured prospectively by remote paper questionnaire between 4 and 10 years after baseline assessment (2015–2016) using the Composite International Diagnostic Interview—Short Form (CIDI-SF) as described previously^2^.

DNAm preprocessing: Genome-wide DNAm data was obtained from whole-blood samples using the Illumina Infinium Methylation EPIC array. Data processing was performed separately for each set. Quality control (QC) and normalisation were conducted using R packages ‘ShinyMethyl’ (version 1.28.0), ‘watermelon’ (version 1.36.0) and ‘meffil’ (version 1.1.1). Details of the protocol are described elsewhere^1^. In summary, quality control procedures removed probes if there was an outlying log median methylated signal intensity against unmethylated signal for each array, or a bead count <3 in ≥5 % of the total probe sample, or a detection p-value >0.05 for set 1 and p-value >0.01 for set 2 in ≥0.5% of the total sample in each respective set. Samples were excluded if sex prediction from methylation data was inconsistent with self-reported data, or a detection p-value >0.05 for set 1 and p-value >0.01 for set 2 found in >1% of the overall probes for each set respectively. The data was then normalised using the ‘dasen’ method from the ‘waterRmelon’ R package (version 1.36.0).

The M-values were corrected using a linear-mixed model, controlling for relatedness using the GCTA-estimated genetic relationship matrix for set 1. This step was omitted for set 2 as all participants were unrelated within the set and to set 1. The residualised M-values for 769,526 autosomal CpG probes were then used for further analysis.

**Avon Longitudinal Study of Parents and Children (ALSPAC) / Accessible Resource for Integrated Epigenomic Studies (ARIES)^3^**

**DOI for Protocol paper:** <https://doi.org/10.1093/bioinformatics/bty476>

MD phenotype: MDD was measured using the Edinburgh Postnatal Depression Scale (EPDS)^4^. Briefly, participants were asked to mark the response closest to how they have been feeling in the past 7 days on a 10-item scale, where the total score is 30 and a score above 13 indicates MDD.23 We transformed the scores into a binary variable, where MDD cases were those who scored above 13 and controls were those with a total score of ≤13.

DNAm preprocessing: Illumina Infinium HumanMethylation450 Beadchip arrays were used for measuring genome-wide DNAm data from peripheral blood samples^3^. The R package ‘meffil’ (version 1.1.1) was used for pre-processing, quality control and normalisation as previously reported^3^. Further removal of probes was conducted based on background detection (p > 0.05) and if they reached beyond the 3 times inter-quantile range from 25 to 75% or identified as cross-hybridising or polymorphic. Related (IBD >0.1) participants were not included in the analyses.

M-value transformation was conducted. In total, 481,600 and 449,595 CpG probes remained for analysis on ALSPAC adults and children, respectively.

**The biological classification of mental disorders (BeCOME)^5^/ The OPtimized Treatment Identification at the MAx Planck Institute (OPTIMA)^6^**

**DOI for Protocol paper:** https://doi.org/[10.1186/s12888-020-02541-z](https://doi.org/10.1186/s12888-020-02541-z) (BeCOME), https://doi.org/[10.1186/s12888-020-02880-x](https://doi-org.emedien.ub.uni-muenchen.de/10.1186/s12888-020-02880-x) (OPTIMA).

Description

The biological classification of mental disorders (BeCOME) cohort is an observational and exploratory study initiated in 2015 by the Max Planck Institute of Psychiatry (MPIP) in Munich, Germany and currently consists of n = 307 participants (16% of which do not meet DIA-X/M-CIDI criteria and are considered healthy controls)^5^. The study was approved by the local Ethics Committee of the Ludwig Maximilians University, Munich, Germany, and written informed consent is obtained from all participants. The study is conducted in accordance with the Declaration of Helsinki.

The OPtimized Treatment Identification at the MAx Planck Institute (OPTIMA) – Trial is a clinical randomized controlled trial (RCT) designed to investigate immediate and long-term effectiveness of Schema Therapy (ST) compared to Individual Supportive Therapy (IST) and Cognitive Behavioral Therapy (CBT) in inpatient and day clinic settings^6^. It consists of 300 depressed patients. The trial is registered at clinicaltrials.gov (NCT03287362). It was approved by the Institutional Ethic Committee of the Faculty of Medicine of LudwigMaximilians-University Munich (project number 17–395). All participants were informed in oral and written form about the study, treatments, assessments and duration of the study. All samples are entered into the biobank at the Max Planck Institute of Psychiatry which has been approved by the ethics committee of the Ludwig-Maximilians-University, Munich, under the project-ID 338–15.

MD phenotype: In OPTIMA and BeCOME, Depression was diagnosed by the M-CIDI^7^, a computerized, fully standardized German version of the WHO-CIDI. For the case-control MWAS, samples from OPTIMA (n = 103, all cases) and BeCOME (n = 119 cases and n = 83 controls) were combined and analysed together. The included MD cases did not meet the criteria for manic episodes, bipolar disorder or schizophrenia.

DNAm preprocessing:

DNA of both cohorts was extracted according to standard procedures and run together on the same arrays. Bisulfite-conversion of DNA was performed with the EZ-96 DNA Methylation kit (Zymo Research, Irvine, CA). Illumina Infinium MethylationEPIC BeadChip (Illumina, San Diego, CA, USA) was used for epigenome-wide methylation analysis according to manufacturer protocols. Preprocessing of methylation data was performed using a standard pipeline^8^ with the *minfi* R package^9^. After loading raw intensity values directly into R version 4.0.5 (R Core Team, 2021) and transforming them into beta-values, a quality control was performed. Samples with a poor detection p value > 0.05, samples presenting with distribution artefacts in raw beta-values or sex mismatches between estimated sex from methylation data and phenotypic sex were excluded. Normalization of beta-values was performed using stratified quantile normalization^10^ and subsequently beta-mixture quantile normalization^11^. Batch effects were sequentially with ComBat of the *sva* R package^12^. Batch-corrected M values were transformed into beta-values and MixupMapper^13^ confirmed that no samples mix-up or swaps during the experiment had taken place.

**BiDirect^14^**

**DOI for Protocol paper:** <https://doi.org/10.1186/1471-244X-14-174>

MD phenotype: All BiDirect participants of the depression cohort were either hospitalised or in outpatient treatment for a depressive episode during recruitment. They received selected modules (i.e. modules A, A’, B, D, and O) of the M.I.N.I. International Neuropsychiatric Interview (German version 5.0.0), which assessed whether a participant exhibited acute (first or recurrent) major depression with or without melancholic features, acute dysthymia, acute/former manic/hypomanic episodes, or acute generalized anxiety disorder. Further, for all patients who received a M.I.N.I. diagnosis of acute major depression, it was clarified whether atypical depression features were currently present by means of six selected items of the Inventory of Depressive Symptomatology (IDS) (items 8, 12, 14, 27, 29, and 30). In addition, all patients with depression were assessed using the 17 items version of the Hamilton Depression Rating Scale (HAM-D-17) and the 14 items version of the Hamilton Anxiety Rating Scale (HAM-A-14) during recruitment and filled in the Centre-for-Epidemiologic-Studies-Depression-Scale (CESD) in the examination. All population controls also filled in the CESD and in case of a score above the established cut-off (>=16) additionally received the above described instruments of the depression cohort^14^.

DNAm preprocessing: Genome-wide DNAm was measured from whole-blood samples collected at the initial visit (baseline) using the Illumina Infinium Methylation EPIC array. The raw data was processed using a custom R pipeline that implements “minfi” for quality control, signal conversion and probe filtering, and “bigmelon” for outlier removal and normalization using “dasen”. Briefly, samples with mean detection p-value > 0.05, as well as probes with detection p-values > 0.01 and those with cross-reactivity (i.e. non-specific), single nucleotide polymorphisms (SNPs) and/or located in sex chromosomes were removed using “minfi”. Consecutively, outlier detection and removal, as well as normalization with the dasen method were performed using “bigmelon”. The normalized beta values were then extracted and converted to M-values. The cell-type composition for blood samples was estimated using “bigmelon”. Principal components analysis was performed with “FactoMineR”. The dataset used for analysis contained 772,867 CpG probes and 591 individuals.

**Biopsychosocial factors of major depression in youth (BioMD-Y/LMU)^15^**

**DOI for Protocol paper:** <https://doi.org/10.1176/appi.ajp.2019.18091014> *.*

Description

The LMU cohort study using the BioMD-Y (“biopsychosocial factors of major depression in youth“) sample is a longitudinal clinical case-control study with the aim to examine which clinical, demographic, (epi-) genetic factors and life events have an influence on the course of depression. Youth between 7 and 18 years of age are recruited since 2009 from two child and adolescent clinics in Munich and the clinic’s website, flyers and local advertisements.  It currently consists of 633 children (n = 381 cases) with available DNA methylation data which were all included in the MWAS. Further characterization of the cohort is described in Scherff et al., (*submitted*). All studies were approved by the local ethics committees and were conducted in accordance with the current version of the Declaration of Helsinki.

MD phenotype: Diagnosis of a current MD episode according to DSM/ ICD is assessed based on the diagnostic interview for mental disorders in children and adolescents (<https://doi.org/10.13154/rub.101.90>), which is administered to the participant and to one parent. The Kinder-DIPS is a well-established, standardized, semi-structured German diagnostic interview (Adornetto et al., 2008; Klin. Diag. Eval., 1, 363–377). Interviewers are psychologists or psychology students who have been specifically trained in administering the Kinder-DIPS and are closely supervised by experienced clinicians. Interrater reliability for depressive disorders on both the parent (Yule’s Q = .71-.94) and child version (Yule’s Q = .71-.94) has been demonstrated (<https://doi.org/10.1026/1616-3443/a000430>).

DNAm preprocessing:

DNA was extracted according to standard procedures. Bisulfite-conversion of DNA was performed with the EZ-96 DNA Methylation kit (Zymo Research, Irvine, CA). Illumina Infinium MethylationEPIC BeadChip (Illumina, San Diego, CA, USA) was used for epigenome-wide methylation analysis according to manufacturer protocols. Preprocessing of methylation data was performed using a standard pipeline (Maksimovic, Phipson, & Oshlack, 2016 with the minfi R package^9^. After loading raw intensity values directly into R version 4.0.5 (R Core Team, 2021) and transforming them into beta-values, a quality control was performed. Samples with a poor detection p value > 0.05, samples presenting with distribution artefacts in raw beta-values or sex mismatches between estimated sex from methylation data and phenotypic sex were excluded. Normalization of beta-values was performed using stratified quantile normalization^10^ and subsequently beta-mixture quantile normalization^11^. Batch effects were sequentially with ComBat of the ‘sva’ R package^12^. Batch-corrected M values were transformed into beta-values and MixupMapper^13^ confirmed that no samples mix-up or swaps during the experiment had taken place.

**Dublin cohort^16^**

**DOI for Protocol paper:** https://doi.org/[10.1093/ecco-jcc/jjv176](https://doi.org/10.1093/ecco-jcc/jjv176)

MD phenotype: MD status was measured using the Beck Depression Inventory (BDI). The BDI is a self-reported scale used to assess severity of depression. The recommended cut-off for clinical depression in medical populations is ≥16 ([https://doi.org/10.1037/t00742-000](https://psycnet.apa.org/doi/10.1037/t00742-000)) was used in this study. We transformed the scores into a binary variable, where MD cases were those who scored 16 and above and controls were those with a total score of <16.

DNAm preprocessing: DNA methylation was quantified using the Infinium HumanMethylation450 BeadChip (Illumina, Inc., San Diego, CA) from peripheral blood mononuclear cells as previously described^16^. GenomeStudio software (Illumina, Inc.) was used to extract signal intensities for each probe and perform initial quality control. Further quality control checks, quantile normalization and separate background adjustment of methylated and unmethylated intensities of type I and II probes were undertaken using the dasen function in the R ‘wateRmelon’ package (available from [www.bioconductor.org](http://www.bioconductor.org/))^17^. Samples with 1% of sites with a detection p value <0.01 or a bead count <3 in 1% of samples (n = 2) were removed from the analysis. Non-specific probes and probes with known single-nucleotide polymorphisms ±10 base pairs (bp) from the single base extension site were removed^18,19^. The final analyses included 430 821 probes, and 186 samples passed our stringent quality control filter. Probes on the X and Y chromosomes were used to confirm the sex of the sample. R package “wateRmelon” was used to compute beta values. The beta2m function from the lumi R package (<https://www.rdocumentation.org/packages/lumi/versions/2.24.0/topics/beta2m>) was used to compute M Values from beta values. Batch effects were controlled for using the ComBAT function^20^ in the ChAMP R package^21^.

**Environmental Risk (E-Risk) Longitudinal Twin Study^22^**

**DOI for Protocol paper:** [**https://doi.org/10.1176/appi.ajp.2017.17060693**](https://doi.org/10.1176/appi.ajp.2017.17060693)

MD phenotype: At age 18, E-Risk Study participants were privately interviewed about their depressive symptoms over the past year using the Diagnostic Interview Schedule (<https://doi.org/10.1002/9780470479216.corpsy0273>). The interview began with four screening questions to identify participants who had experienced at least 2 weeks of persistent low mood, anhedonia, or irritability in the past year, or those who had been prescribed medication for depression. Participants who answered positively to any of the screening items were asked a further 24 questions designed to map onto the nine symptom-criteria of a major depressive episode specified in the DSM-IV. We created a scale based on the total number of symptom-criteria present. To identify participants with clinically significant depression we used a diagnostic cut-off based on the presence of at least five symptom-criteria plus interference in daily functioning. At age 18, 20% of E-Risk Study participants met these criteria for MDD.

DNAm preprocessing: DNA methylation was assayed from blood samples collected at the age 18 time-point, using the Illumina Infinium HumanMethylation450 BeadChip. A total of 1669 samples (out of 1700 available whole blood samples) were assayed; 31 samples were not useable (e.g., due to low DNA concentration). Approximately 500ng of DNA from each sample was treated with sodium bisulfite, using the EZ-96 DNA Methylation kit (Zymo Research, CA, USA). DNA methylation was quantified using the Illumina Infinium HumanMethylation450 BeadChip (Illumina Inc, CA, USA) (“Illumina 450K array”) run on an Illumina iScan System (Illumina, CA, USA) using the manufacturers’ standard protocol. Members of complete twin pairs (n = 743 pairs; n = 1486 individuals) were randomly assigned to bisulfite-conversion plates and Illumina 450K arrays, with siblings processed in adjacent positions to minimise batch effects.

Data from 1669 unique individuals entered the Quality Control (QC) pipeline, which was performed in the R statistical programming environment. Data were imported using the ‘methylumIDAT’ function from the methylumi package. The first QC step assessed the signal intensities, excluding samples with both median methylated (‘M’) and unmethylated (‘U’) intensities <2500 (n = 10). Second, the fully methylated controls were identified - their intensity profiles were characteristic of the sample being fully methylated and confirmed that no plate rotations or plate mislabelling had occurred – and subsequently removed from the dataset. Third, using ten control probes included on the 450K array, we examined the efficiency of the sodium bisulfite conversion reaction; samples (n = 1) were excluded if their “conversion score” was <80. Fourth, multidimensional scaling was performed for DNA methylation probes on each of the sex chromosomes and compared to the reported gender; we identified a discrepancy for 2 samples from 2 different twin pairs. As both individuals were part of MZ twin pairs, we compared the genotypes of 65 SNP probes for both members of each twin pair separately and observed that they were not genetically identical to their reported twin. The samples were genetically identical to the other’s twin pair. As the unique identifiers for these individuals were rearrangements of the same 4 numbers, we concluded that these discrepancies were a result of a sample switch at the lab. Correcting the sample ID, and repeating the gender checks as previously described, led to a 100% match with the corresponding phenotype information so no sample was excluded. Fifth, to confirm genetic identity of the DNA samples, we assessed genotype concordance between SNP probes on the 450K array and data generated using Illumina OmniExpress24v1.2 genotyping BeadChips. Data were available for 35 of the 65 SNP probes from both platforms for 1638 (98.7%) of the samples. 1658 samples passed the stringent QC pipeline. The data were then processed with the ‘pfilter’ function from the wateRmelon package^17^ excluding 0 samples with >1% of sites with a detection p value > 0.05, 567 sites with beadcount < 3 in 5% of samples and 1448 probes with >1% of samples with detection p value > 0.05. The data were normalised with the dasen() function from the wateRmelon package. Prior to any analyses, probes with common (> 5% MAF) SNPs within 10 bp of the single base extension and probes with sequences previously identified as potentially hybridising to multiple genomic loci were excluded (n = 52,760)^18,19^. After QC and annotation of CpGs to the 450k array 430,802 sites remained for analysis. Methylation beta-values were transformed to M-values using logit transformation.

These Illumina DNA methylation data are accessible from the Gene Expression Omnibus (accession code: GSE105018).

**EXTEND^23^**

MD phenotype: MD status was defined as a binary variable based on self-report. Specifically, participants were identified as having MD if they answered ‘yes’ to the question: "Has a doctor ever diagnosed you with depression requiring regular medical treatment?".

DNAm profiling: DNA methylation data for the EXTEND cohort were generated using the Illumina Infinium HumanMethylationEPIC BeadChip array (‘EPIC array’). The EZ-96 DNA Methylation-Gold kit (Zymo Research; Cat No# D5007) was used for treating 500 ng of DNA from each sample with sodium bisulfite. Raw data were processed using the wateRmelon package^17^ and put through a stringent quality control pipeline including the following steps: (1) checking methylated and unmethylated signal intensities and excluding poorly performing samples; (2) assessing the chemistry of the experiment by calculating a bisulphite conversion statistic for each sample, excluding samples with a conversion rate <80%; (3) identifying the fully methylated control sample was in the correct location; (4) multidimensional scaling of sites on the X and Y chromosomes separately to confirm reported sex; (5) using the 59 SNPs on the Illumina EPIC array to check for sample duplications; (6) use of the ‘pfilter’ function in wateRmelon to exclude samples with >1% of probes with a detection *P*-value > 0.05 and probes with >1% of samples with detection *P*-value > 0.05; (7) normalization of the DNA methylation data was performed using the ‘dasen’ function in wateRmelon^17^; (8) samples that were dramatically altered as a result of normalization were excluded on the basis of the difference between the normalized and raw data; and (9) removal of cross-hybridizing and SNP probes^24,25^.

**Finnish Twin Register^26,27^**

MD phenotype: In two parts of the Finnish Twin Cohort (FinnTwin12^26^ and FinnTwin16 Study^27^), depression was assessed among twins during in-person studies  of selected subsamples. The young adult participants were interviewed using the Semi-Structured Assessment for Genetics of Alcohol (SSAGA)^28^ psychiatric interviews. The depression phenotype was defined according to the criteria of the DSM-IV major depressive disorder (MDD) diagnosis (APA, 1994, <https://doi.org/10.1176/ajp.152.8.1228>).

DNAm preprocessing: Blood DNAm was quantified using the Infinium HumanMethylation450 or EPIC BeadChip Kit (Illumina, San Diego, CA, USA) according to the manufacturer’s protocol. Methylation data was preprocessed using R package *meffil*^3^. First, beta values were background corrected and then functional normalization was performed, using 13 PCs for the 450k dataset and 17 PCs for the EPIC dataset. Probes with a detection p-value > 0.05 or a beadcount < 3 in greater than 10% of samples were removed. Beta-mixture quantile normalization was used to adjust beta values for differences because of probe type^11^. Methylation beta values were transformed to M-values using beta2m function in the *lumi* package in R. We estimated cell types using the FlowSorted.Blood.EPIC reference set. After QC, 1,351 samples with good quality 450K or EPIC data remained for analysis.

**FOR2107^29^**

MD phenotype: Lifetime MDD was assessed using a semi-structured interview according to DSM-IV-TR (Diagnostic and Statistical Manual of Mental Disorders) (Wittchen et al., 1997, <https://doi.org/10.1026//0084-5345.28.1.68>) applied by trained staff.

DNAm preprocessing: DNA methylation was profiled in peripheral blood using the Illumina Infinium MethylationEPIC BeadChip. During quality control with the minfi and ewastools packages in R, samples were excluded in case of technical issues detected via Illumina control probes, a call rate <98%, sex mismatch, a mismatch in SNP fingerprints or relatedness. Methylation probes were excluded in case of a call rate <98%, SNPs inside the probe body or a non-cg type. After stratified quantile normalisation, M values were extracted for downstream analyses. A list of probes flagged by McCartney et al. (2016) as cross-reactive or polymorphic with an allele frequency >5% in the European population (n=52,541) was prepared for exclusion during downstream analyses.

**Janssen^30^**

**DOI for Protocol paper:** <https://pubmed.ncbi.nlm.nih.gov/36319817/>

Cohort Description**:** A total of 191 blood samples from 112 patients with MDD were collected up till the interim analysis from an observational clinical study OBSERVEMDD0001 (ClinicalTrials.gov Identifier: NCT02489305), where a patient must have met DSM-V criteria for nonpsychotic, recurrent MDD within the past 24 months (ie, the start of the most recent major depressive episode (MDE) must be ≤ 24 months before screening); have a Montgomery Asberg Depression Rating Scale (MADRS) total score ≤ 14 at screening and baseline visits; have evidence of recent response (within the past 3 months) to an oral antidepressant treatment regimen (taken at an optimal dosage and for an adequate duration, and be currently taking and responding to an oral antidepressant treatment regimen. The samples from the participants with MDD could have been obtained from either a baseline visit or a follow-up visit. 32 samples from 32 healthy controls self-reported to be free of MDD were collected by BioIVT and used as control samples for this Cohort. The institutional review boards of all participating clinical trial sites reviewed and approved the study and patients provided informed consent for DNA sample collection.

The OBSERVEMDD0001 study was approved by the respective local or central Institutional Review Boards (IRBs) overseeing the clinical sites participating in the study, these included the University of Pennsylvania Office of Regulatory Affairs IRB, University of Iowa IRB, Baylor College Of Medicine IRB, University of Michigan IRB, University of Cincinnati IRB, Sharp HealthCare IRB, Springfield Committee for Research Involving Human Subjects (SCRIHS), Western IRB, University of Kansas School of Medicine—Wichita Human Subjects Committee,

Rush University Medical Center IRB, Hartford Hospital IRB, University of Massachusetts Medical School IRB, and Sterling IRB. In addition, the BioIVT samples were collected with IRB approval from Schulman IRB. All clinical studies and sample collections were carried out following the ethical principles outlined in the Declaration of Helsinki and are consistent with Good Clinical Practices and applicable regulatory requirements. All patients provided written informed consent before entry into the study.

DNAm data generation and preprocessing: Whole blood samples were collected, and DNA was extracted for methylation profiling. DNA methylation was measured using Infinium® MethylationEPIC BeadChip (Illumina, Inc., San Diego, CA, USA) at 850 000 CpG sites throughout the genome. The assay for the cohort (both cases and controls) was performed in one batch. Quality control of the EPIC array data was performed using R package ChAMP. Probes with detection p value ≥ 0.01 in one or more samples (n = 14,421), or with bead count less than 3 in at least 5% of samples (n = 8999), non-CG probes (n = 2625), probes with known SNP sites or with cross-reactivity (n = 93,722), probes align to multiple locations on the genome (n = 15), as well as probes located on the sex chromosomes (n = 16,532) were filtered out. The methylation levels were normalized using the Dasen method in the R package wateRmelon. The blood cell composition was estimated using the estimateCellCounts function in minfi which used a reference blood dataset of fluorescence-activated cell sorting (FACS) sorted CD8T, CD4T, NK, B cell, monocytes, granulocytes, and eosinophils. Surrogate variables are covariates inferred from high-dimensional data that are used in subsequent analyses to adjust for unknown and/or unmodeled sources of noise. We used R package sva (v3.38) to estimate surrogate variables for unknown sources of variation to remove artefacts in the epigenetic profile experiments. Removing batch effects using surrogate variables before downstream differential analysis has been shown to improve reproducibility.

**Munich Antidepressant Response Signature (MARS/GSK)^31,32^**

**DOI for Protocol paper:** https://doi.org/[10.1016/j.jpsychires.2008.05.002](https://doi.org/10.1016/j.jpsychires.2008.05.002) (MARS) https://doi.org/[10.1093/hmg/ddl166](https://doi.org/10.1093/hmg/ddl166) (UniDep)

Cohort Description

The Munich Antidepressant Response Signature (MARS) project^32^ is a naturalistic longitudinal clinical study providing a representative of depressed inpatients (n = 842) from Southern Bavaria, Germany. Baseline data was collected between 1995 and 2005. The study was approved by the local Ethics Committee of the Ludwig Maximilians University, Munich, Germany, and carried out in accordance with the latest version of the Declaration of Helsinki.

The Unipolar Depression study (UniDep)^31^ is a cross-sectional case-control study. It consists of German patients (n = 1000) with recurrent MDD from in- and outpatients at the Max-Planck-Institute of Psychiatry in Munich and psychiatric hospitals in Augsburg and Ingolstadt, located close to Munich. Healthy controls (n = 1029) were randomly selected from a Munich-based community sample, screened for presence of anxiety and affective disorders using the Composite International Diagnostic Screener (CIDI), and matched for ethnicity, age, and sex (to 5-year intervals). Baseline data was collected between 2002 and 2004. Further characterization of the cohort is described in Lucae et al., (2006). The study was approved by the Ethics Committee of the Ludwig Maximilians University in Munich, Germany, and written informed consent was obtained from all subjects.

**MD phenotype**: MDD status in MARS was determined by a trained physician upon hospitalization and confirmed by the Munich-composite international Diagnostic Interview (M-CIDI) in 2009.  UniDep patients were diagnosed by WHO-certified raters according to the DSM-IV using the Schedule for Clinical Assessment in Neuropsychiatry (SCAN). UniDep healthy controls were screened for presence of anxiety and affective disorders using the Composite International Diagnostic Screener (CIDI). For the case-control MWAS, samples from MARS (n = 199, all cases) and UniDep (n = 112 cases and n = 186 controls) were combined and analyzed together.

DNAm preprocessing:

Genomic DNA from both cohorts was extracted from peripheral blood using the Gentra Puregene Blood Kit (Qiagen) and run together on the same arrays.  To minimize batch effects, samples were randomized with respect to case-control status, sex and age. Genomic DNA was bisulfite converted using the Zymo EZ-96 DNA Methylation Kit (Zymo Research) and DNA methylation levels were assessed for >480,000 CpG sites using the Illumina HumanMethylation450 BeadChip array. Hybridization and processing were performed according to the instructions of the manufacturer.

The Bioconductor R package *minfi* (version 1.10.2)^9^ was used for the quality control of methylation data including intensity read outs, normalization, cell type composition estimation, *β*-value and *M*-value calculation. Outliers, i.e., samples whose behavior deviated from that of others in terms of median intensity, were excluded from the analysis (*N* = 3 in the discovery sample, *N* = 5 in the replication sample) as well as samples with a discordant methylation-predicted vs. reported sex (*N* = 1 in the replication sample). Failed probes were excluded based on a detection *P*-value larger than 0.01 in >50% of the samples. X and Y chromosome were removed to avoid a possible gender effect and also non-specific binding probes^18^. We also excluded probes if single nucleotide polymorphisms (SNPs) were documented in the interval for which the Illumina probe is designed to hybridize. Probes located close (10 bp from query site) to a SNP which had a minor allele frequency of ≥0.05, as reported in the 1000 Genomes Project, were also removed. This yielded a total of around 425,000 CpG sites in the discovery and replication sample for further analysis.The data were then normalized with functional normalization (FunNorm)^33^, an extension of quantile normalization included in the R package *minfi*.

Batch effects were identified by inspecting the association of principal components of the methylation levels with possible technical batches using linear regressions and visual inspection of principal component analysis plots using the Bioconductor R package *shinyMethyl* (version 0.99.3). Identified batch effects (i.e., bisulfite conversion plate and plate position) were removed using the Empirical Bayes’ method *ComBat*^20^. Batch corrected *M*-values after *ComBat* were used for all further statistical analyses.

**Lothian Birth Cohort 1936 (LBC1936)^34^**

**DOI for Protocol paper:** <https://doi.org/10.1186/s13073-018-0585-7>

DNAm in LBC1936 was profiled in whole blood samples using the HumanMethylation450 BeadChip Kit, Ilumina, San Diego, CA, USA^35^. Details on sample preparation and quality check have been reported previously^36^. Briefly, background correction was performed, and quality control was utilised to remove probes that had a low detection rate (p > 0.01 for more than 5% of samples), low call rate (p < 0.01 for < 95% of probes) and low quality (manual inspection); and to remove samples that had a poor match between genotypes and single nucleotide polymorphism (SNP) control probes, and incorrectly predicted sex.

R function “*beta2m*” from the “*lumi*” package^37^ was used for M-value transformation. M-values were corrected for effects of sample plate, BeadChip, position on BeadChip, and estimated cell type proportions (CD4^+^ T cells, CD8^+^ T cells, B cells, Natural Killer Cells and granulocytes), and residuals from this model were used for the methylation score calculation. Methylation principal components (PCs) were calculated based on M-values residualised on age, sex, and the technical and biological variables noted above. Cross-reactive and polymorphic CpGs (N= 41,709), identified by Chen et al. (2013) were removed, resulting in 417,600 CpGs across the 22 autosomes^18^.

**Netherlands Twin Register (NTR)^38^**

**DOI for Protocol paper:** <https://doi.org/10.1038/ncomms11115>

Cohort description: The Netherlands Twin Register (NTR) is a population-based cohort of over 200,000 twins and twin-families from across the Netherlands. NTR respondents are periodically invited to participate in lab and questionnaire measurements, resulting in a vast array of biological and behavioural data. A detailed description of NTR participants and data waves is available elsewhere^39^. Good quality whole blood DNA methylation data were available for 3087 samples from 3055 individuals, including monozygotic and dizygotic twins, parents of twins, siblings of twins and spouses of twins. For 32 individuals, longitudinal methylation data were available (two time points, mean range=5.2 year). The major depression MWAS included N = 2771 individuals.

DNA methylation: DNA was collected from buccal cells and whole blood as part of multiple projects. For the current paper, we analysed DNA methylation measured in whole blood collected in the NTR-Biobank study^38,40,41^. DNA methylation (DNAm) was measured with the Infinium HumanMethylation450 BeadChip Kit (Illumina, San Diego, CA, USA) by the Human Genotyping facility (HugeF) of ErasmusMC, the Netherlands (http://www.glimdna.org/) as part of the Biobank-based Integrative Omics Study (BIOS) consortium^42^. DNA methylation measurements have been described previously (Bonder et al., 2017; van Dongen et al., 2016). Genomic DNA (500ng) from whole blood was bisulfite treated using the Zymo EZ DNA Methylation kit (Zymo Research Corp, Irvine, CA, USA), and 4 µl of bisulfite-converted DNA was measured on the Illumina 450k array following the manufacturer’s protocol. A number of sample- and probe-level quality checks and sample identity checks were performed, as described in detail previously (van Dongen et al., 2016). In short, sample-level QC was performed using MethylAid^43^. Probes were set to missing in a sample if they had an intensity value of exactly zero, or a detection p > .01, or a bead count of < 3. After these steps, probes that failed based on the above criteria in >5% of the samples were excluded from all samples (only probes with a success rate ≥ 0.95 were retained). The following probes were also removed: sex chromosomes, probes with a single nucleotide polymorphism (SNP) within the CpG site (at the C or G position) irrespective of minor allele frequency in the Genome of the Netherlands (GoNL) population, irrespective of minor allele frequency^44^, and ambiguous mapping probes reported by Chen et al with an overlap of at least 47 bases per probe^18^. DNAm data were normalized with functional normalization^33^.

Major depression: Lifetime major depressive disorder (MDD) data were derived from the Composite International Diagnostic Interview (CIDI) (Kessler, Wittchen, et al., 1998, <https://doi.org/10.1002/mpr.33>), the Lifetime Depression Assessment Self-report (LIDAS)^45^, and DSM-oriented Adult Self Report scales (ASR) (Achenbach & Rescorla, 2003) of the Achenbach System of Empirically Based Assessment (ASEBA). The Dutch computerized version of the CIDI was administered in a telephone interview in 1997 and 2007 in order to obtain lifetime MDD status according to diagnostic criteria of the DSM-4^46^. The LIDAS was developed as part of the Biobanking and Biomolecular Resources Research Infrastructure (BBMRI-NL) to efficiently identify lifetime MDD in population-based cohorts, and was distributed in 2015-2020 among NTR respondents^47^. LIDAS is based on the Composite International Diagnostic Interview short form (CIDI-sf) (Kessler, Andrews, et al., 1998, <https://doi.org/10.1002/mpr.47>) and contains diagnostic self-report items to determine lifetime MDD status in accordance with DSM-5 criteria. DSM-oriented scale scores of the ASR were derived from longitudinal NTR surveys (1991-2013), containing 10 to 14 items with ratings on a 3-point scale. Within-survey scale scores were converted to z scores to account for variation in the number of items, and subsequently converted to T scores, where a threshold of 69 was applied to identify MDD cases (> 69) and controls (< 69). Data from the CIDI, LIDAS and ASR scales were combined to form one numeric variable coded as 1 (lifetime MDD case) / 0 (lifetime MDD control). When data from multiple sources were available for an individual, the information from CIDI was favoured over that of LIDAS and ASR, and data from LIDAS over that of ASR.

Data availability: Data are available from the Netherlands Twin Register upon reasonable request (<https://tweelingenregister.vu.nl/information_for_researchers/working-with-ntr-data>, accessed on 24 August 2023).

**Study of Health in Pomerania (SHIP)^48^**

**DOI for Protocol paper:** https://doi.org/10.1186/s13059-015-0600-x

Cohort description: The Study of Health in Pomerania (SHIP) is a longitudinal general population-based study designed to assess the prevalence of common risk factors and diseases in a population living in Western Pomerania, Germany^49^. The SHIP-Trend sub-cohort comprises of 8,016 adults randomly drawn from local registers in September 2008 with the data collection of 4,420 subjects aged 20 to 84 concluding in September 2012.

The SHIP study is conducted in accordance with the Helsinki Declaration and was reviewed and approved by the Institutional Review Board of the University Medicine Greifswald. All subjects provided informed written consent.

MD phenotype: Major depression (MD, lifetime) has been determined based on DSM-IV criteria (Diagnostic and Statistical Manual of Mental Disorders IV (Wittchen 1994, Wittchen, Lachner et al. 1998, Wittchen, Höfler et al. 1999, American Psychiatric Association 2000).

DNA methylation processing: DNA was extracted from blood samples of n=508 SHIP-Trend participants to assess DNA methylation using the Illumina HumanMethylationEPIC BeadChip array. Samples were randomly selected based on availability of multiple OMICS data, excluding type II diabetes, and enriched for prevalent myocardial infarction. The samples were taken between 07:00 AM and 04:00 PM, and serum aliquots were prepared for immediate analysis and for storage at -80 °C in the Integrated Research Biobank (Liconic, Liechtenstein). Processing of the DNA samples was performed at the Helmholtz Zentrum München. Preparation and normalization of the array data was performed according to the CPACOR workflow^48^ using the software package R ([www.r-project.org](http://www.r-project.org)). The array idat files were processed using the minfi package. Probes that had a detection p-value above background (sum of per-array methylated and unmethylated intensity values based p-value ≥1E-16) were set to missing. Methylation beta values were calculated as proportion of methylated intensity value on the sum of methylated+unmethylated+100 intensities. Arrays with observed technical problems (±4 SD outside control probe intensity mean) during steps like bisulfite conversion, hybridization or extension, as well as arrays with mismatch between sex of the proband and sex determined by the chr X and Y probe intensities were removed from subsequent analyses. Additionally, only arrays with a call rate ≥ 95% were processed further resulting in 495 samples with methylation data on 865,859 sites available for subsequent analyses.

To account for potential confounding effects due to blood cell composition, blood cell subtypes were estimated by the Houseman method^50^ and included in the association model. Additionally, the array processing batch (n=248 and n=247) and the first six principal components of the control probe intensities obtained by the CPACOR workflow were included in the model to account for technical factors.

Data availability: Further information on the SHIP data used in the preparation of this article can be found here: https://fvcm.med.uni-greifswald.de/dd_service/data_use_intro.php. Qualified researchers can request access to SHIP data through a research proposal at the following link: <https://fvcm.med.uni-greifswald.de/cm_antrag/index.php>.

**TwinsUK**^51^

**DOI for Protocol paper:** <http://doi.org/10.1017/thg.2019.65> (DNA methylation); <https://wellcomeopenresearch.org/articles/4-10> (self-reported depression)

MD phenotype and covariates: Depression status was ascertaining using self-reported responses to a series of questionnaires administered to participants between 2004 and 2017. Three different variations of the question were asked; “Have you ever had depression diagnosed by GP or psychiatrist?”, “Have you ever been told by a doctor or other health professional that you had; clinical depression”, “Have you ever suffered depression (depression means feeling low, lacking interest in life and not enjoying things you usually enjoy for at least 2 weeks)?”. We defined a composite measure of “nearest” depression status by taking the response to a depression question nearest to the date of each individual’s DNA methylation sample collection date. Altogether, 937 individuals with DNA methylation data had at least one response to a depression question. We excluded participants who did not have a response within 5 years of the DNA methylation sample collection. The majority of depression status data in the remaining subset of twins were derived from responses within one year of DNA methylation sample collection, and 94.5% of depression data derived from questionnaire responses were within 2 years of DNA methylation sample collection.

Alcohol consumption (average grams of alcohol consumed per day) and smoking status were self-reported Using the nearest year the data entry in relation to the whole blood sample collection^52,53^.

BMI was calculated using the nearest available measurement of height and weight to the time of DNA methylation as Kg/m^2^.

DNAm preprocessing: This study used up to 692 whole blood samples from individuals in the TwinsUK cohort. Whole blood DNA methylation levels were assessed using the Illumina Infinium HumanMehtylation450 BeadChip array, as previously described^51^. Quality Control (QC) was performed using ENmix^54^ and minfi^9^ in the R statistical programming environment^55^ Briefly, minfi was used to exclude samples with median methylated and unmethylated signals below 10.5. ENmix was used for general QC and excluding probes and samples that did not meet standard parameter values. ENmix was further used for background and dye bias correction (bgParaEst=”oob”), quantile normalization of signals (method=”quantile1”), and estimation of adjusted β-values (with ‘rcp’). Signals with detP > 0.000001 and nbead < 3 were set to ‘NA’, and cross-reactive or multimapping probes were removed as previously described^51^. After QC, 430,802 sites remained for downstream analysis.

Statistical analysis: Methylation beta values were converted to M-values using logit transformation. MWAS were conducted using the lmrse package in R^56^, which accounts for relatedness amongst twin pairs using a linear model with cluster robust standard errors. All samples were female. In model 1, age, zygosity (MZ/DZ), estimated cell composition (CD8T, CD4T, NK, Bcell, Mono, Gran derived from the Houseman algorithm^50^) and 20 surrogate variables (SVs) calculated using the SmartSVA package^57^, were included as covariates. Model 2 adjusted additional for smoking status by way of a proxy, the *AHRR* CpG cg05575921. Model 3 adjusted additionally for BMI and alcohol consumption. Individuals with missing values for any of the covariates were excluded from the analyses. Following exclusions, the resulting sample size was N=692 (201 cases and 491 controls) in model 1 and model 2, and N=652 (193 cases and 459 controls) in model 3.

**Understanding Society/ UK Household Longitudinal Study (UKHLS)^58^**

**DOI for Protocol paper:** <https://doi.org/10.1016/j.ajhg.2018.09.007>

MD phenotype: For the basic and smoking-adjusted models, dataset 1 contains 1121 individuals from the British Household Panel Survey (BHPS) component of Understanding Society (72 cases) and dataset 2 contains 2361 individuals (242 from BHPS and 2119 from the General Population Sample (GPS)) (277 cases). For the complex model, dataset 1 contains 693 individuals (39 cases) and dataset 2 contains 1562 individuals (148 cases). MD phenotype inclusion criteria are: 1) Must have at least one non-missing value across the 16 depression-related variables; 2) Self-reported sex at blood collection is concordant with our main survey sex variable and genotyped sex; and 3) Covariate data is all non-missing.

Eight timepoints of two questions were used regarding seeking for professional health care for depression (variables ‘hcond17’ and ‘hcondn17’ in <https://www.understandingsociety.ac.uk/documentation/mainstage/dataset-documentation/variable/hcond17>). Anyone who responded “yes” to at least one of these questions at any wave was assigned a phenotype of “1”; anyone who had at least one non-missing response and never responded “yes” to any of these questions was assigned a phenotype of “0”.

DNAm preprocessing: Methylation data was in M-value format. Un-residualised M-values were used. Related individuals were excluded by calculating pairwise PI-HAT from our genetic data, filtering for pairs with a value greater than 0.2 and removing individuals based on the column with the lowest number of unique individuals. Probes were removed according to the three lists in McCartney et al^24^.

**Taiwan Biobank**

The Taiwan Biobank (TWB) is a prospective, population-based biobank that recruits adult individuals aged 30 and above. Since October 2012, the TWB has successfully enrolled over 173K community-based volunteers who were free from prior cancer diagnoses. These volunteers contributed DNA and serum samples for the purpose of genotyping and biochemistry assessments. 63.5% of the participants in the TWB were female. Given Taiwan's predominantly Han Chinese population, the public TWB data release is limited to individuals of Han Chinese ethnicity.

The participants underwent physical examinations and engaged in structured interviews that covered demographics, lifestyle behaviours, environmental exposures, dietary patterns, family medical history, and other health-related details documented within a questionnaire. Self-reported depression diagnosis was also recorded.

From the total participant pool, a subset of 2,309 individuals was randomly chosen for DNA methylation (DNAm) quantification analysis. This involved scrutinizing blood DNAm levels using the Illumina Infinium MethylationEPIC BeadChip (Illumina, Inc., San Diego, CA), a platform encompassing around 860,000 CpG sites.

**Supplementary results for MWAS in East Asian ancestry**

We inspected the EWAS Atlas and GWAS Catalog for the top 10 CpGs and annotated genes. Pathway analysis was conducted on those CpGs that met a significance threshold of p <1x10^-5^ (N_CpG_=24).

The mean age in the TBB was 50 years (SD=11 years), with females representing 50% and 61% of the healthy control and case samples, respectively. The top CpG identified in TBB was cg26665746 (β=-0.107, p=2x10^-7^). The gene it is annotated to (EFCC1) has previously been linked to platelet count in European and East Asian ancestries^59^ (Supplementary Table 4).

There were no significant pathways after FDR correction (GO minimum p=7.4x10^-4^: positive regulation of respiratory burst involved in inflammatory response; KEGG minimum p=0.005: neomycin, kanamycin, and gentamicin biosynthesis). Top GO terms and KEGG pathways are listed in Supplementary Table 5.

**Supplementary Figure 1.** Distribution of standard errors for individual studies.

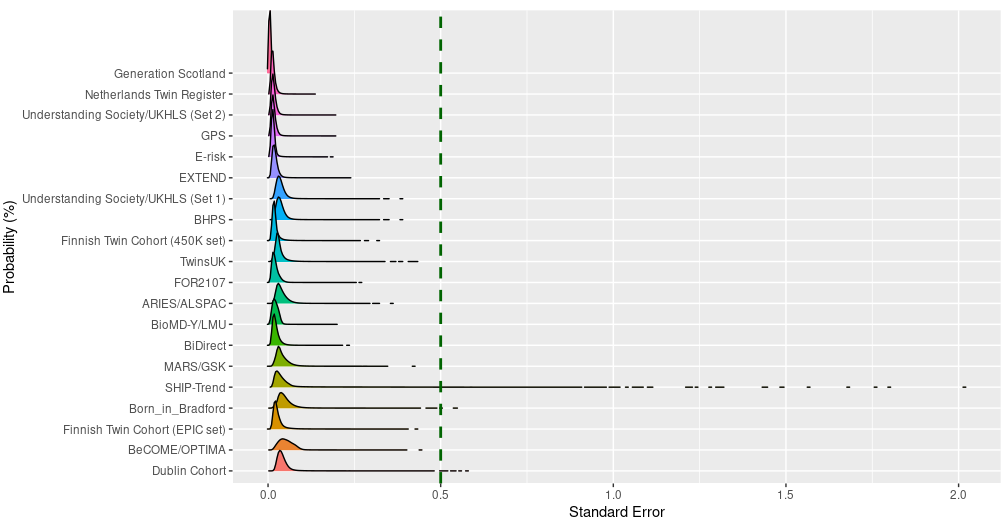

**Supplementary Figure 2.** Sample size of each individual study for the complex model (BMI and alcohol consumption were included as additional covariates).

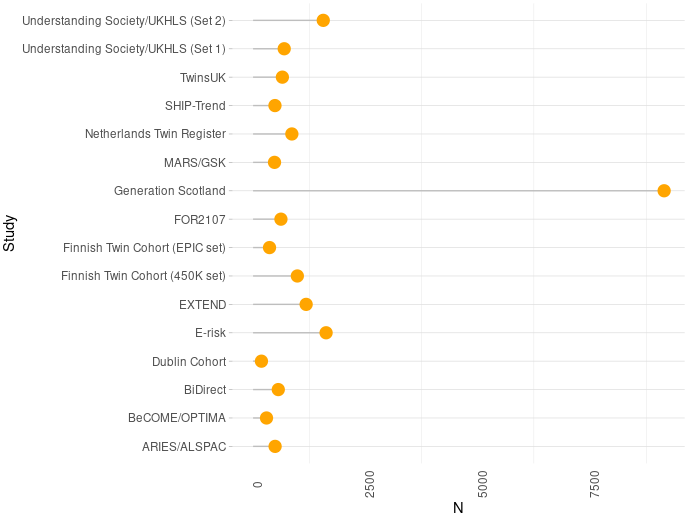

**Supplementary Figure 3.** Inflation factor for summary statistics for each individual study.

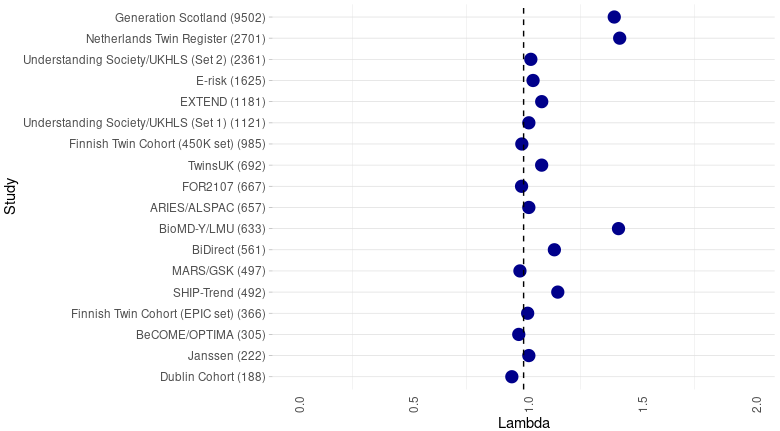

**Supplementary Figure 4**. Effect sizes of each individual study for the significant CpG sites.

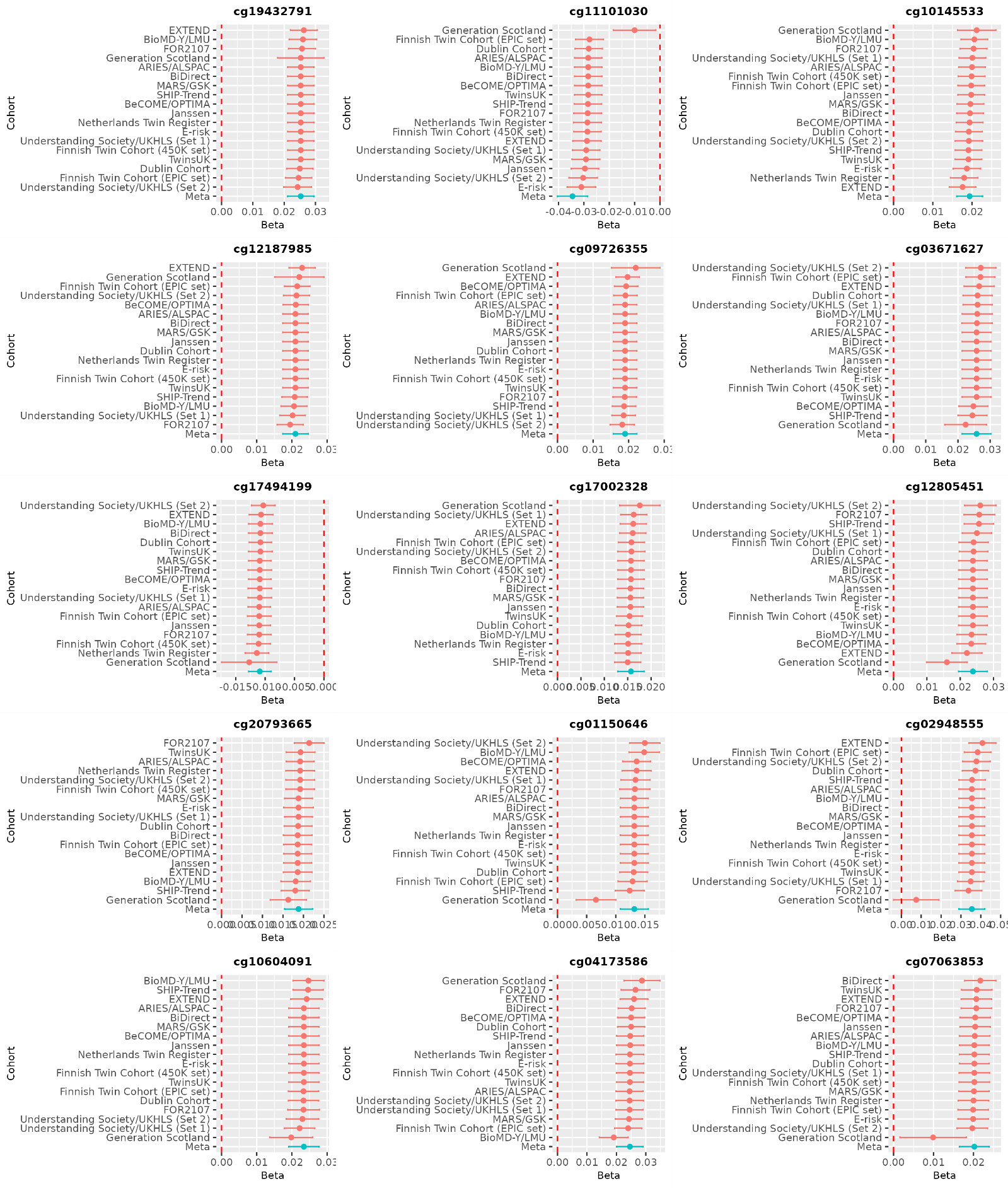

**Supplementary Figure 5**. Proteome-wide association analysis for DNAm score for MD.

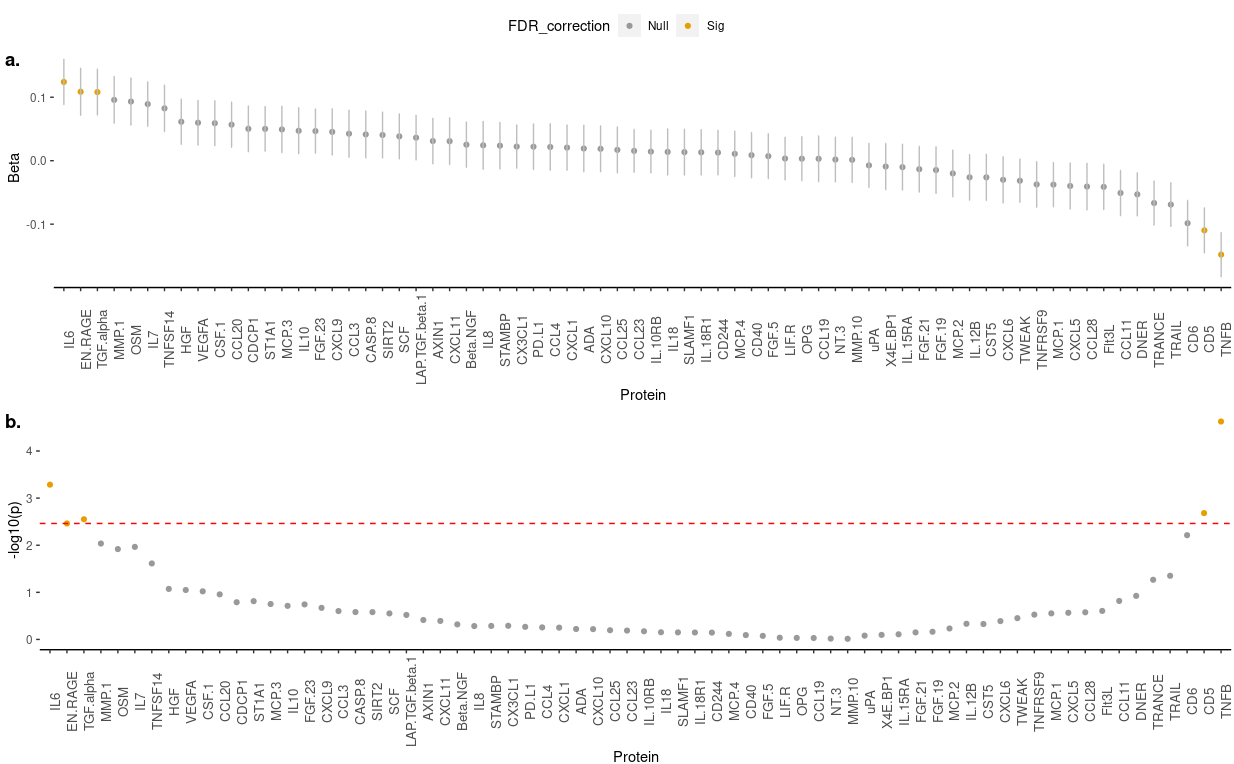

**Supplementary Figure 6.** Forest plot for leave-one-out meta-analysis.

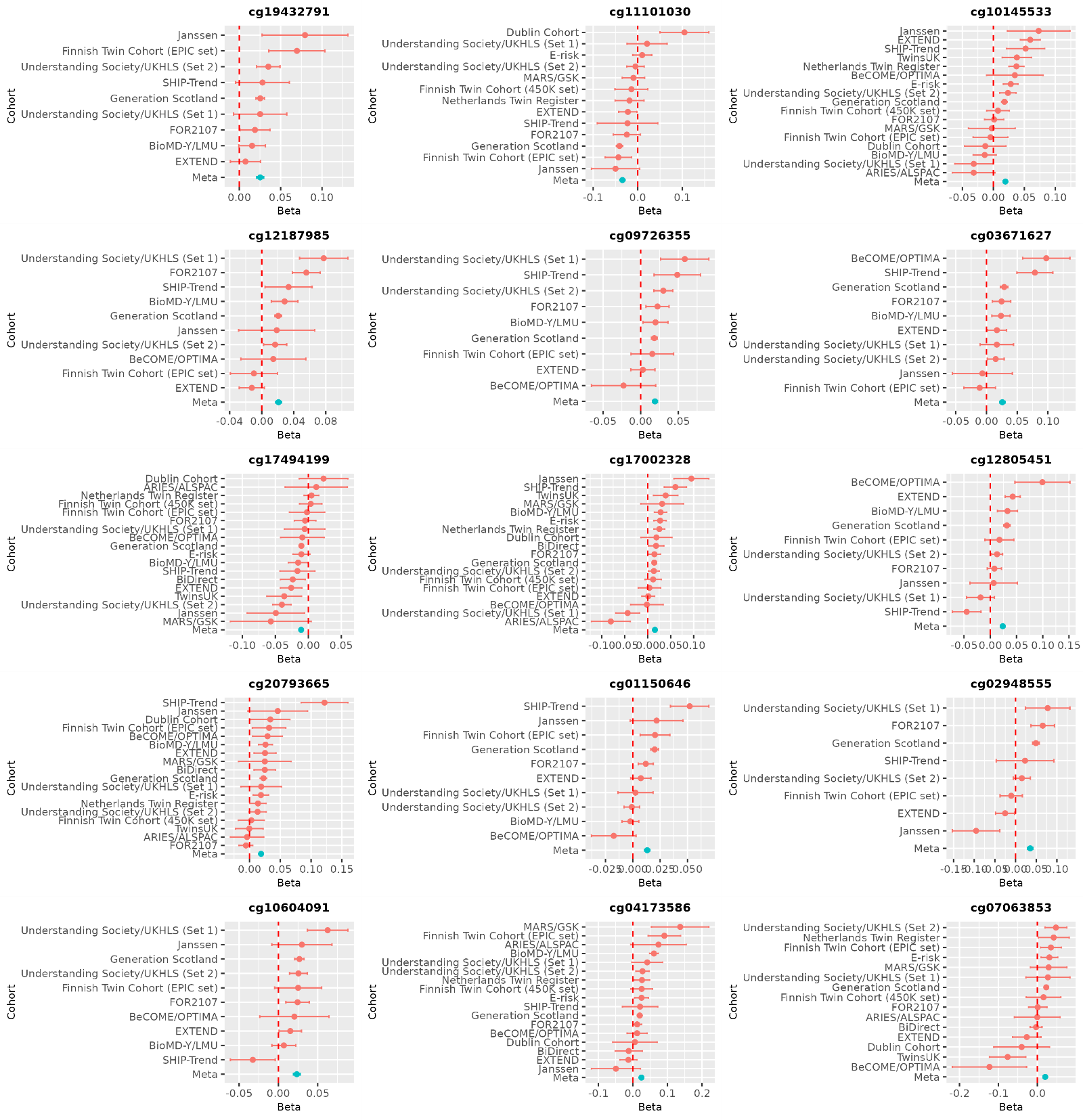

**Supplementary Figure 7.** Meta-regression looking at association between mean age of a study and effect size for MDD MWAS.

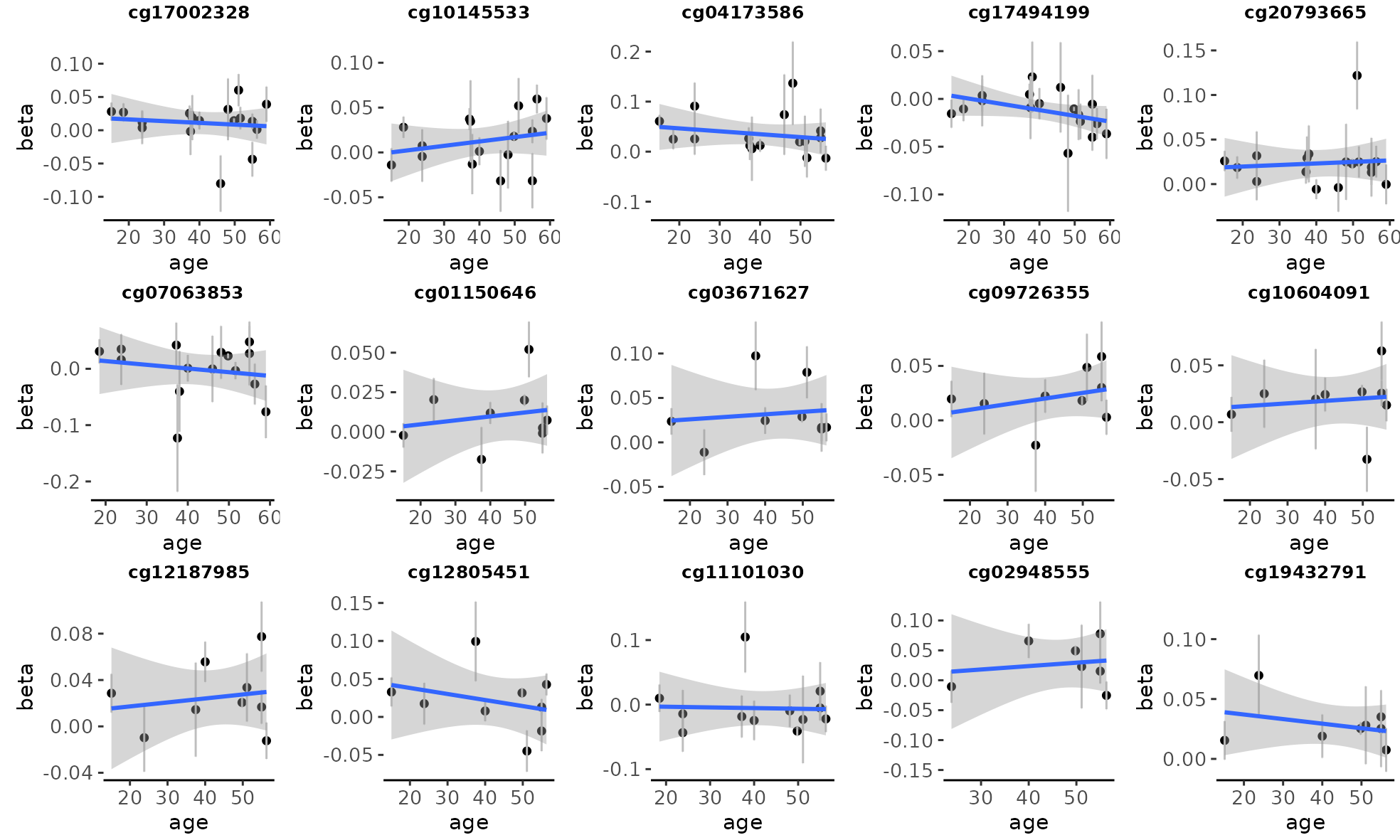

**Supplementary Table 1.** Significant differentially methylated regions (DMRs).

| CHR | DMR start | DMR end | Genes | N_CpG_ | Beta | SE | p | P_Bonferroni_ |
| --- | --- | --- | --- | --- | --- | --- | --- | --- |
| 6 | 30853258 | 30854233 | DDR1 | 12 | 0.036 | 0.005 | 9.89E-13 | 6.75E-08 |
| 19 | 37825211 | 37825455 | HKR1 | 8 | 0.036 | 0.005 | 5.66E-12 | 3.87E-07 |
| 10 | 135342560 | 135343248 | CYP2E1 | 5 | 0.031 | 0.005 | 1.42E-10 | 9.68E-06 |
| 3 | 18480242 | 18481064 | SATB1 | 8 | 0.048 | 0.009 | 1.16E-07 | 0.007921 |
| 6 | 30698784 | 30698936 | FLOT1 | 4 | 0.046 | 0.009 | 1.45E-07 | 0.009885 |
| 11 | 43333495 | 43333512 | API5 | 3 | 0.055 | 0.009 | 2.59E-09 | 1.77E-04 |
| 17 | 7832769 | 7832932 | KCNAB3 | 3 | 0.036 | 0.005 | 1.14E-13 | 7.78E-09 |
| 9 | 139557250 | 139557920 | EGFL7 | 4 | -0.059 | 0.011 | 2.20E-08 | 0.001501 |
| 2 | 219738714 | 219738732 | WNT6 | 2 | 0.074 | 0.014 | 5.44E-08 | 0.003711 |
| 3 | 11643427 | 11643630 | VGLL4 | 3 | 0.083 | 0.015 | 4.45E-08 | 0.003035 |
| 8 | 28479277 | 28480107 |  | 6 | 0.029 | 0.005 | 7.73E-08 | 0.005279 |
| 14 | 81426015 | 81426577 | TSHR | 3 | 0.054 | 0.010 | 1.63E-07 | 0.011122 |
| 17 | 79128827 | 79129078 | AATK | 6 | -0.048 | 0.009 | 2.42E-08 | 0.001652 |
| 5 | 78985495 | 78985588 | CMYA5 | 3 | 0.046 | 0.007 | 2.51E-11 | 1.71E-06 |
| 12 | 120763716 | 120763869 | PLA2G1B | 3 | 0.095 | 0.014 | 1.98E-12 | 1.35E-07 |
| 14 | 50065065 | 50065270 | PPIL5 | 6 | 0.035 | 0.007 | 2.30E-07 | 0.015735 |
| 14 | 91751773 | 91752093 | CCDC88C | 2 | 0.086 | 0.015 | 2.66E-09 | 1.82E-04 |
| 19 | 19221392 | 19221572 | SLC25A42 | 2 | 0.053 | 0.010 | 3.07E-07 | 0.020964 |
| 1 | 153599671 | 153600156 | S100A13;S100A1 | 6 | 0.008 | 0.001 | 2.44E-07 | 0.016632 |
| 2 | 115419537 | 115420053 | DPP10 | 5 | -0.048 | 0.009 | 3.54E-07 | 0.0242 |
| 5 | 131409352 | 131409637 | CSF2 | 4 | -0.039 | 0.007 | 1.25E-07 | 0.008507 |
| 8 | 101315498 | 101315560 | RNF19A | 4 | -0.067 | 0.013 | 1.78E-07 | 0.012131 |
| 2 | 179344387 | 179344870 | FKBP7;PLEKHA3;MIR548N | 3 | 0.060 | 0.012 | 2.34E-07 | 0.016002 |
| 4 | 15779642 | 15779729 | CD38 | 3 | 0.053 | 0.010 | 3.87E-07 | 0.026441 |
| 4 | 153273933 | 153274105 | FBXW7 | 2 | 0.089 | 0.017 | 1.68E-07 | 0.011471 |
| 5 | 43483764 | 43483906 | C5orf28 | 2 | 0.041 | 0.008 | 1.92E-07 | 0.013093 |
| 6 | 31127004 | 31127357 | CCHCR1;TCF19 | 7 | 0.043 | 0.008 | 2.44E-08 | 0.001663 |
| 6 | 31698109 | 31698223 | DDAH2 | 4 | 0.036 | 0.007 | 5.91E-07 | 0.040339 |
| 11 | 61659669 | 61659716 | FADS3 | 2 | 0.070 | 0.013 | 2.18E-07 | 0.014909 |
| 12 | 24104007 | 24104094 | SOX5 | 2 | 0.081 | 0.016 | 4.78E-07 | 0.032654 |
| 6 | 31275643 | 31275791 |  | 6 | -0.038 | 0.006 | 1.30E-11 | 8.91E-07 |
| 4 | 174098862 | 174099006 | GALNT7 | 2 | 0.114 | 0.022 | 2.38E-07 | 0.016214 |
| 11 | 64107517 | 64107672 | CCDC88B | 2 | 0.051 | 0.010 | 6.18E-08 | 0.004221 |
| 12 | 131590460 | 131590596 | GPR133 | 2 | 0.066 | 0.012 | 2.15E-08 | 0.001471 |
| 13 | 51482542 | 51482592 | RNASEH2B;RNASEH2B-AS1 | 2 | 0.090 | 0.017 | 1.64E-07 | 0.011202 |
| 15 | 75660209 | 75660389 | MAN2C1 | 4 | 0.040 | 0.008 | 2.44E-07 | 0.016652 |
| 16 | 85577361 | 85577847 |  | 3 | -0.046 | 0.009 | 3.50E-07 | 0.02391 |

**Supplementary Table 2.** Top 10 results of gene ontology and KEGG pathway enrichment analysis

|  | ONTOLOGY | TERM | ID | N | DE | P | P.FDR |
| --- | --- | --- | --- | --- | --- | --- | --- |
| Gene ontology | BP | Negative regulation of protein localization to ciliary membrane | GO:1903568 | 1 | 1 | 8.14E-04 | 1 |
|  | BP | 1-phosphatidyl-1d-myo-inositol 3,5-bisphosphate metabolic process | GO:1903100 | 1 | 1 | 9.06E-04 | 1 |
|  | BP | Regulation of protein localization to ciliary membrane | GO:1903567 | 2 | 1 | 9.75E-04 | 1 |
|  | MF | Histone methyltransferase activity (h3-k79 specific) | GO:0031151 | 1 | 1 | 1.18E-03 | 1 |
|  | BP | Phosphatidylinositol 5-phosphate metabolic process | GO:1904562 | 2 | 1 | 1.74E-03 | 1 |
|  | MF | Bbsome binding | GO:0062063 | 3 | 1 | 2.43E-03 | 1 |
|  | BP | Negative regulation of protein localization to cilium | GO:1903565 | 3 | 1 | 2.50E-03 | 1 |
|  | BP | Protein targeting to membrane | GO:0006612 | 120 | 2 | 2.79E-03 | 1 |
|  | MF | 1-phosphatidylinositol-3-phosphate 5-kinase activity | GO:0000285 | 3 | 1 | 2.88E-03 | 1 |
|  | MF | 1-phosphatidylinositol-5-kinase activity | GO:0052810 | 3 | 1 | 2.88E-03 | 1 |
| KEGG pathway | -- | Transcriptional misregulation in cancer | hsa05202 | 183 | 2 | 0.007 | 1 |
|  | -- | Lysine degradation | hsa00310 | 61 | 1 | 0.047 | 1 |
|  | -- | Inositol phosphate metabolism | hsa00562 | 69 | 1 | 0.057 | 1 |
|  | -- | Phosphatidylinositol signaling system | hsa04070 | 92 | 1 | 0.074 | 1 |
|  | -- | Phagosome | hsa04145 | 143 | 1 | 0.082 | 1 |
|  | -- | Pathogenic escherichia coli infection | hsa05130 | 189 | 1 | 0.121 | 1 |
|  | -- | Motor proteins | hsa04814 | 191 | 1 | 0.128 | 1 |
|  | -- | Wnt signaling pathway | hsa04310 | 165 | 1 | 0.129 | 1 |
|  | -- | Regulation of actin cytoskeleton | hsa04810 | 218 | 1 | 0.157 | 1 |
|  | -- | Metabolic pathways | hsa01100 | 1464 | 2 | 0.235 | 1 |

**Supplementary Table 3**. Meta-regression looking at association between mean age per study and effect sizes.

| CpG | Beta | SE | df | p |
| --- | --- | --- | --- | --- |
| cg17002328 | -3.16E-04 | 2.73E-04 | 15 | 0.265 |
| cg10145533 | 5.05E-04 | 4.07E-04 | 14 | 0.234 |
| cg04173586 | -9.25E-04 | 2.87E-04 | 14 | 0.006 |
| cg17494199 | -3.48E-04 | 2.14E-04 | 15 | 0.125 |
| cg20793665 | -1.11E-06 | 2.78E-04 | 15 | 0.997 |
| cg07063853 | -6.09E-04 | 5.62E-04 | 13 | 0.298 |
| cg01150646 | 1.99E-04 | 3.65E-04 | 7 | 0.603 |
| cg03671627 | 1.20E-04 | 4.64E-04 | 7 | 0.804 |
| cg09726355 | 6.66E-05 | 3.60E-04 | 7 | 0.858 |
| cg10604091 | 3.66E-04 | 4.00E-04 | 7 | 0.39 |
| cg12187985 | -1.17E-04 | 6.49E-04 | 7 | 0.862 |
| cg12805451 | -3.54E-04 | 7.31E-04 | 7 | 0.643 |
| cg11101030 | -4.12E-04 | 6.28E-04 | 10 | 0.527 |
| cg02948555 | 2.87E-04 | 0.001 | 5 | 0.846 |
| cg19432791 | 6.73E-05 | 3.31E-04 | 6 | 0.846 |

**Supplementary Table 4. Results in the Taiwan Biobank for the significant CpG sites found in the meta-MWAS of the European samples.**

| CpG | Taiwan Biobank | | | | European-ancestry meta-analysis | | |  |
| --- | --- | --- | --- | --- | --- | --- | --- | --- |
|  | **Beta** | **SE** | **p** | **N** | **Beta** | **SE** | **p** | **N** |
| cg01150646 | 0.013 | 0.011 | 0.251 | 2072 | 0.013 | 0.002 | 3.80E-08 | 16850 |
| cg02948555 | 0.027 | 0.035 | 0.427 | 2072 | 0.036 | 0.007 | 4.72E-08 | 15911 |
| cg03671627 | 0.025 | 0.019 | 0.191 | 2072 | 0.026 | 0.005 | 9.85E-09 | 16850 |
| cg04173586 | 0.022 | 0.030 | 0.46 | 2072 | 0.025 | 0.005 | 6.30E-08 | 24044 |
| cg07063853 | -0.047 | 0.071 | 0.506 | 2072 | 0.020 | 0.004 | 6.42E-08 | 23396 |
| cg09726355 | -0.001 | 0.021 | 0.96 | 2072 | 0.019 | 0.003 | 8.53E-09 | 16628 |
| cg10145533 | -0.017 | 0.020 | 0.381 | 2072 | 0.019 | 0.003 | 5.34E-09 | 24192 |
| cg10604091 | -0.009 | 0.020 | 0.633 | 2072 | 0.023 | 0.004 | 5.27E-08 | 16850 |
| cg11101030 | -0.035 | 0.033 | 0.29 | 2072 | -0.035 | 0.006 | 4.99E-09 | 15901 |
| cg12187985 | 0.026 | 0.022 | 0.237 | 2072 | 0.021 | 0.004 | 5.43E-09 | 16850 |
| cg12805451 | 0.006 | 0.019 | 0.75 | 2072 | 0.024 | 0.004 | 3.11E-08 | 16850 |
| cg17002328 | 0.029 | 0.017 | 0.094 | 2072 | 0.016 | 0.003 | 2.32E-08 | 24751 |
| cg17494199 | -0.002 | 0.018 | 0.919 | 2072 | -0.011 | 0.002 | 1.22E-08 | 24749 |
| cg19432791 | 0.005 | 0.020 | 0.802 | 2072 | 0.025 | 0.004 | 1.70E-09 | 16545 |
| cg20793665 | 8.00E-04 | 0.023 | 0.972 | 2072 | 0.019 | 0.003 | 3.79E-08 | 24753 |

**Supplementary Table 5**. Top 10 CpGs identified in the Taiwan Biobank cohort. Background information for each CpG and gene was extracted from EWAS Atlas (<https://ngdc.cncb.ac.cn/ewas/atlas>) and GWAS (<https://www.ebi.ac.uk/gwas/>) catalogue databases. All associations included in the table from these two catalogues are genome-wide significant. EUR=European ancestry; EA=East Asian; SA=South Asian; H/L=Hispanic/Latin American; AFR=African American; AFC=Afro-Caribbean.

| CpG site | Gene | CHR | Beta | SE | P | CpG look-up | Gene look-up |
| --- | --- | --- | --- | --- | --- | --- | --- |
| cg26665746 | EFCC1 | 3 | -0.1073 | 0.0205 | 1.83E-07 |  | Platelet count (EUR; EA; SA; AFR; AFC), BMI-adjusted whr (EUR)^60,61^ |
| cg13127506 | CDK2AP1 | 12 | -0.1195 | 0.0242 | 8.22E-07 | Obesity^62^ | Apolipoprotein A1 levels (EUR), Educational attainment (EUR), Mathematical ability (EUR)^63,64^ |
| cg08506585 |  | 13 | -0.1874 | 0.0379 | 8.35E-07 | Fetal alcohol spectrum disorder^65^ |  |
| cg11178592 | HK1 | 10 | -0.1262 | 0.0256 | 9.09E-07 |  | Hemoglobin A1 levels (EUR; H/L; EA; AFR; AFC), Hematocrit (EUR; H/L; AFR; AFC), Mean corpuscular volume (EUR; H/L; AFR; AFC)^66,67^ |
| cg17840132 | DAB2 | 5 | 0.0939 | 0.0196 | 1.73E-06 |  | Creatinine (EUR; EA), Heel bone mineral density (EUR), Height (EUR; EA)^66,67^ |
| cg19208791 | CFAP54 | 12 | -0.0854 | 0.0178 | 1.76E-06 |  | Adolescent idiopathic scoliosis^68^ |
| cg23363804 | USP34 | 2 | 0.1441 | 0.0301 | 1.86E-06 |  | Mean corpuscular volume (EUR), Brain measurement (EUR), Educational attainment (EUR)^64,69^ |
| cg09895934 |  | 21 | -0.0943 | 0.0198 | 1.92E-06 |  |  |
| cg13755233 | LOC101927378 | 7 | 0.1322 | 0.0284 | 3.36E-06 |  |  |
| cg23010118 |  | 6 | -0.1403 | 0.0301 | 3.39E-06 |  |  |

**Supplementary Table 6**. Pathway analysis conducted on TBB CpGs using p < 1x10^-5^ (N=24). The two resources utilise are Gene Ontology (GO) and Kyoto Encyclopaedia of Genes and Genomes (KEGG). Highlighted rows are those overlapping with the European ancestry MWAS meta-analysis (N=3). N=number of genes in the term; DE=number of genes that are differentially methylated; P.DE=p-value for over-representation of the term; FDR=false discovery rate.

|  | ONTOLOGY | TERM | ID | N | DE | P | P.FDR |
| --- | --- | --- | --- | --- | --- | --- | --- |
| Gene ontology | BP | Positive Regulation Of Respiratory Burst Involved In Inflammatory Response | GO:0060265 | 2 | 1 | 7.24E-04 | 1 |
|  | BP | Leading Edge Cell Differentiation | GO:0035026 | 1 | 1 | 1.13E-03 | 1 |
|  | BP | Transition Between Slow And Fast Fiber | GO:0014886 | 1 | 1 | 1.61E-03 | 1 |
|  | BP | Urea Homeostasis | GO:0097274 | 2 | 1 | 1.88E-03 | 1 |
|  | BP | Negative Regulation Of Respiratory Burst Involved In Inflammatory Response | GO:0060266 | 5 | 1 | 2.77E-03 | 1 |
|  | BP | Negative Regulation Of Respiratory Burst | GO:0060268 | 5 | 1 | 2.77E-03 | 1 |
|  | BP | Regulation Of Respiratory Burst Involved In Inflammatory Response | GO:0060264 | 6 | 1 | 3.09E-03 | 1 |
|  | BP | Respiratory Burst Involved In Inflammatory Response | GO:0002536 | 7 | 1 | 3.57E-03 | 1 |
|  | BP | Nucleolus Organization | GO:0007000 | 6 | 1 | 0.004 | 1 |
|  | BP | Regulation Of Cellular Hyperosmotic Salinity Response | GO:1900069 | 4 | 1 | 0.004 | 1 |
| KEGG pathway | - | Neomycin, Kanamycin And Gentamicin Biosynthesis | path:hsa00524 | 5 | 1 | 0.005 | 1 |
|  | - | Galactose Metabolism | path:hsa00052 | 30 | 1 | 0.022 | 1 |
|  | - | Starch And Sucrose Metabolism | path:hsa00500 | 32 | 1 | 0.022 | 1 |
|  | - | Fructose And Mannose Metabolism | path:hsa00051 | 33 | 1 | 0.024 | 1 |
|  | - | Basal Transcription Factors | path:hsa03022 | 44 | 1 | 0.027 | 1 |
|  | - | Biosynthesis Of Nucleotide Sugars | path:hsa01250 | 37 | 1 | 0.027 | 1 |
|  | - | Amino Sugar And Nucleotide Sugar Metabolism | path:hsa00520 | 49 | 1 | 0.035 | 1 |
|  | - | Carbohydrate Digestion And Absorption | path:hsa04973 | 43 | 1 | 0.035 | 1 |
|  | - | Glycolysis / Gluconeogenesis | path:hsa00010 | 67 | 1 | 0.041 | 1 |
|  | - | Type Ii Diabetes Mellitus | path:hsa04930 | 46 | 1 | 0.046 | 1 |
